# Probability of causation in individual workers: Lung cancer due to occupational exposure to asbestos

**DOI:** 10.64898/2026.02.06.26345596

**Authors:** Javier Mancilla-Galindo, Susan Peters, Huifang Deng, Henk van der Molen, Hans Kromhout, Lützen Portengen, Roel Vermeulen, Dick Heederik

**Affiliations:** Institute for Risk Assessment Sciences, Utrecht University, Utrecht, the Netherlands; Department of Public and Occupational Health, Amsterdam UMC, Amsterdam, the Netherlands; National Expertise Centre for Substance-related Occupational Diseases (Lexces), Utrecht, the Netherlands

**Keywords:** Asbestos, Lung cancer, Probability of Causation, Causality, Workers’ Compensation

## Abstract

**Background:** Compensation systems for asbestos-related lung cancer commonly apply the Helsinki criteria, which assume a 4% excess relative risk per fibre-year. Under linearity, the relative risk (RR) doubles at 25 fibre-years, where the probability of causation (PoC), estimated as (RR−1)/ RR, reaches 50%. Recent large case-control studies suggest steeper exposure-response relations at lower exposures. We aimed to estimate PoC of asbestos-related lung cancer to evaluate exposure thresholds for compensation of lung cancer cases occupationally exposed to asbestos.

**Methods:** Relative risk of asbestos-related lung cancer was estimated using two approaches:

- A meta-regression of 22 occupational studies forming the core evidence on cumulative asbestos exposure and lung cancer since the 1980s (130,341 participants).
- A meta-analysis of the recently conducted SYNERGY pooled case-control study (14 studies, 37,866 participants), adjusted for age, sex, smoking, and study.

PoC was estimated with 95% prediction intervals (95%PI) and interpreted as a lower bound.

**Results:** Occupational cohort studies showed a shallow non-linear exposure-response relation at low exposure ranges (0–65 fibre-years) with substantial heterogeneity (I2 = 92.7%). SYNERGY showed a steeper relation (6.8% increase in risk per fibre-year; 95%PI: 0%–17.5%) and lower heterogeneity (I2 = 63.4%). PoC ≥50% occurred at 62.9 (point estimate) and 18.2 fibre-years (upper 95%PI) for occupational studies, versus 10.5 and 4.3, respectively, in SYNERGY.

**Conclusions:** The SYNERGY study, a high-quality study with harmonised exposure assessment, provided exposure-response estimates relevant to current exposure to lower levels of mixed asbestos fibres in the Netherlands, supporting lower exposure thresholds than the existing Helsinki criteria when estimating PoC in compensation contexts.

**Key Messages:** *What is already known on this topic:*

- Probability of causation (PoC) is applied in workers’ compensation with a PoC ≥50% as criterion, typically corresponding to a doubling in baseline disease risk;
- An exposure threshold of 25 fibre-years is used as the doubling in risk criterion for lung cancer attribution (2014 Helsinki criteria) in compensation systems worldwide;
- Upper confidence intervals (statistical uncertainty) of the PoC have been suggested as eligibility thresholds for compensation to weigh uncertainties in favour of workers.

*What this study adds:*

- This study uses recent and transparent quantitative meta-analyses for the relation between lung cancer and asbestos exposure as alternative for the Helsinki criterion of 25 fibre-years (4% excess relative risk per fibre-year);
- This study applies the upper bound of 95% prediction intervals of relative risk to estimate PoC, considering uncertainty because of between study differences in risk, which could be a more appropriate measure than statistical uncertainty of the meta-risk-estimate alone;
- Together with the use of recent general population studies, a lower exposure threshold (4.3 fibre-years) was obtained than when the Helsinki criterion was applied (25 fibre-years);
- In contrast, historical occupational cohort studies resulted in a threshold of 18.2 fibre-years.

*How this study might affect research, practice or policy:*

- Prediction intervals are a potentially useful tool to account for uncertainty arising from meta-analyses in compensation processes;
- Caution is warranted when interpreting PoC as it does not account for earlier cancer onset due to exposure. Future research could aim to address this gap.

## Introduction

Assessing whether diseases are caused by occupational exposures often occurs within national workers’ compensation systems. Determining causality between occupational exposure and disease requires integrating epidemiological, experimental animal, and mechanistic evidence, including toxicological findings.^1^ Population-based epidemiological studies are particularly important as human evidence is critical for classifying agents as proven human carcinogens. Lung cancer, a high impact disease with individual and societal burden, has multiple causes, with cigarette smoking being the leading risk factor.^2^ However, various occupational exposures also substantially increase lung cancer risk, including asbestos, respirable crystalline silica, diesel engine exhaust, arsenic, nickel, chromium, among others which are classified as respiratory carcinogens.^2,3^ For multi-causal diseases like lung cancer, different causes produce clinically and histologically indistinguishable tumours, so individual aetiology cannot be determined, for instance, from tumour features alone. Despite this, compensation schemes commonly require establishing criteria for assessment of individual claims.

A common approach is to estimate the likelihood of disease occurrence attributable to occupational exposure. This has been operationalised as the probability of causation (PoC), expressed as (RR−1)/RR, where RR is the disease risk ratio comparing exposed to non-exposed, defined for a specific risk period. When PoC ≥50%, the disease is presumed “more likely than not” caused by the exposure. This approach has been applied in various societal contexts including tort law and cancer compensation due to radiation,^4–6^ coal tar pitch volatiles,^7^ smoking,^8^ among others. It has been shown that PoC is generally not identifiable from epidemiological data alone since exposure may accelerate disease onset regardless of whether disease ultimately occurs, and background risk is typically heterogeneous across individuals.^9,10^ In this sense, it is argued that (RR−1)/RR represents a lower bound of the true PoC.^9^

Strict interpretations of PoC require causal assumptions for its estimation from epidemiological studies,^11^ such as specifying a biological model,^12^ that exposure cannot prevent the disease, and the ability to estimate unbiased risk ratios which are transferable to individuals in whom PoC is applied. Errors arising from limitations in study design, conduct and analysis, such as exposure or disease measurement error and misclassification, confounding, causal model misspecification, and imprecision often impede a causal interpretation of PoC. Therefore, it has been argued that uncertainties should benefit the claimant when applying PoC in occupational compensation contexts.^4,6,7^

In the Netherlands, a one-time compensation scheme for substance-related occupational diseases has been in place since 2022, adopting the *presumably plausible principle (‘voorshands aannemelijk principe’)*, operationalised by considering uncertainties in favour of workers.^13,14^ Two main sources of uncertainty are considered: (1) uncertainties in reconstructing individual cumulative exposure when applying job-exposure matrices,15 and (2) uncertainties in exposure-response.14 Asbestos-related lung cancer was among the first occupational diseases included in January 2022.

Asbestos was completely banned in the Netherlands in 1993 following earlier bans on crocidolite in 1977 and asbestos spraying in 1978. Between 6,800 and 17,500 cases of asbestos-related lung cancer from pre-1993 exposures were projected for 2011 to 2030,^16^ with an estimated 600 new cases occurring in 2023.^17^ In many countries, lifetime cumulative asbestos exposure of 25 fibre-years is considered the threshold for a doubling in lung cancer risk and compensation eligibility, resulting from the 4% excess risk per fibre-year as proposed in the 2014 Helsinki report.^18^ This assumes that the relative risk increases linearly with cumulative exposure *x* (fibre-years): *RR*(*x*) = 1 + 0.04*x*. At 25 fibre-years, *RR* = 1 + 0.04 × 25 = 2, and thus *P oC* = (2 − 1)/2 = 0.5 or 50%. However, this estimate has been criticised for lacking methodological transparency.^19^ Specifically, the 4% value was not based on a formal meta-analysis of all available evidence and assumes a linear exposure-response relation, inconsistent with recent studies indicating that exposure-response may be steeper at lower exposure and levels off at higher exposure.^20^ Moreover, the exposure-response slope is significantly affected by study exposure assessment quality.^20–22^

Sources of error in asbestos exposure assessment include limited detection capability when sampling and analysis of fibres was not yet fully developed, difficulty applying consistent fibre-to-mass or fibre-type conversion factors in case of exposure studies conducted using different measurement techniques,23 and incomplete or imprecise job histories.^21^ Together, these generally result in underestimated or attenuated exposure-response, estimated to be as large as a factor of 1.5 to 2.21,22

The Dutch compensation scheme requires a transparent and reproducible PoC estimate for occupational lung cancer due to asbestos. In the absence of a nationally representative study with the necessary exposure and confounder data to estimate PoC, a meta-analysis of the published literature was considered. Thus, this study aimed to estimate the PoC of asbestos-related lung cancer, to evaluate exposure thresholds for financial compensation of lung cancer cases occupationally exposed to asbestos, treating (RR−1)/RR as a conservative lower bound on the PoC, and weighing uncertainties to favour claimants. We estimated exposure levels associated with a doubling in lung cancer risk (PoC ≥ 50%) based on two series of studies:

- A meta-regression of studies with quantitative exposure-response relations, recently conducted as part of a systematic review for occupational standard setting in Europe by the European Chemicals Agency (ECHA). The study base consisted of 20 occupational cohorts and two case-control studies providing excess risk estimates for asbestos cumulative exposure categories to different types of asbestos in mining and production industries, including cohorts with mixed exposure. In most studies, information on cigarette smoking was not available.
- The study base created by ECHA contained a meta-analysis of a multi-country general population pooled case-control study, the SYNERGY study, which was also analysed separately as the only study with homogeneous exposure assessment across individual studies, covering populations recruited more recently, from a wide range of occupational settings, with lower levels of exposure to mixed asbestos fibres, and data on important confounders (e.g., smoking).

## Methods

### Study design and setting

#### Occupational asbestos studies

Occupational asbestos studies were most recently evaluated by the European Chemicals Agency (ECHA) using the systematic review by van der Bij, et al.,^20^ an update of the initial review by Lenters, et al.,^21^ which considered 19 epidemiological studies. ECHA updated risk assessment to derive an occupational exposure standard in 2021 by updating^24–26^ or adding studies,^27–29^ including SYNERGY,^26^ a pooled case-control study described below. Characteristics of occupational studies are presented in **Table 1**. The occupational cohort studies were conducted in different countries, in earlier periods than SYNERGY, and mainly involve mining and specific asbestos production industries, using different asbestos types depending on product requirements. Each study had its exposure assessment component (**Supplementary Table 1**), with most estimating workers’ lifetime cumulative asbestos exposure using job exposure matrices (JEM), and further categorising by cumulative exposure to estimate excess lung cancer risk. Only two studies reported smoking-adjusted relative risks.^26,28^ Because these studies started in different periods and exposed populations were followed over several decades, different techniques were used between and within studies to measure asbestos: counting dust particles (e.g., impinger) in earlier periods and, more recently, quantifying fibres (e.g., phase contrast microscopy). This often required applying particle-to-fibre conversion factors between and within studies, known to have significant error.^23^ Therefore, applying a single conversion factor to whole blocks of person-time may shift a study’s entire exposure axis multiplicatively, biasing its exposure-response slope in study-specific directions rather than only reducing precision. Accordingly, studies were rated by Lenters, et al.^21^ and ECHA^22^ for exposure assessment quality (low/high) based on documentation, exposure contrast, conversion factors (internal/external), measurement period coverage relative to population exposure, and job history sufficiency.

**Table 1.**
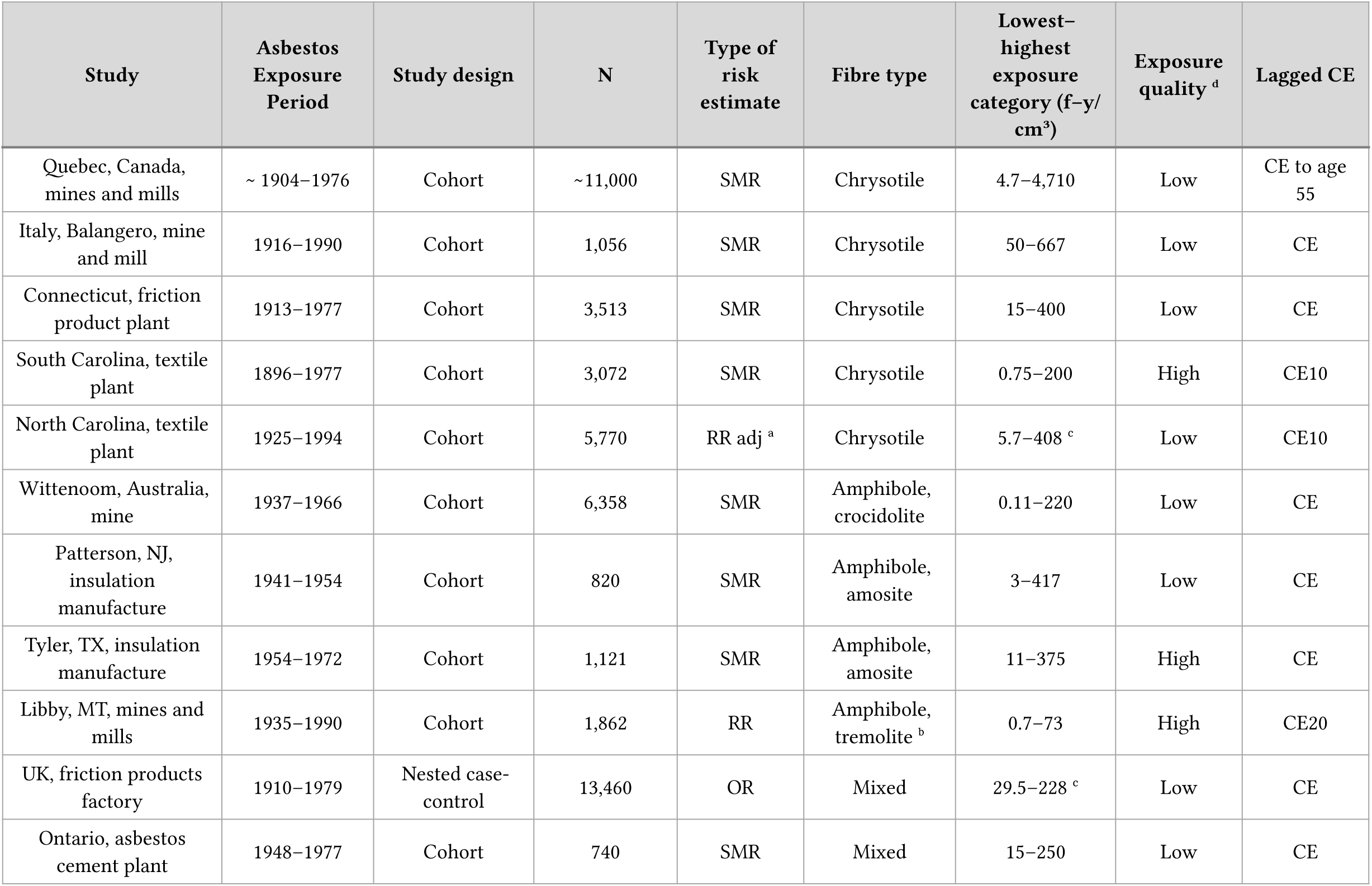
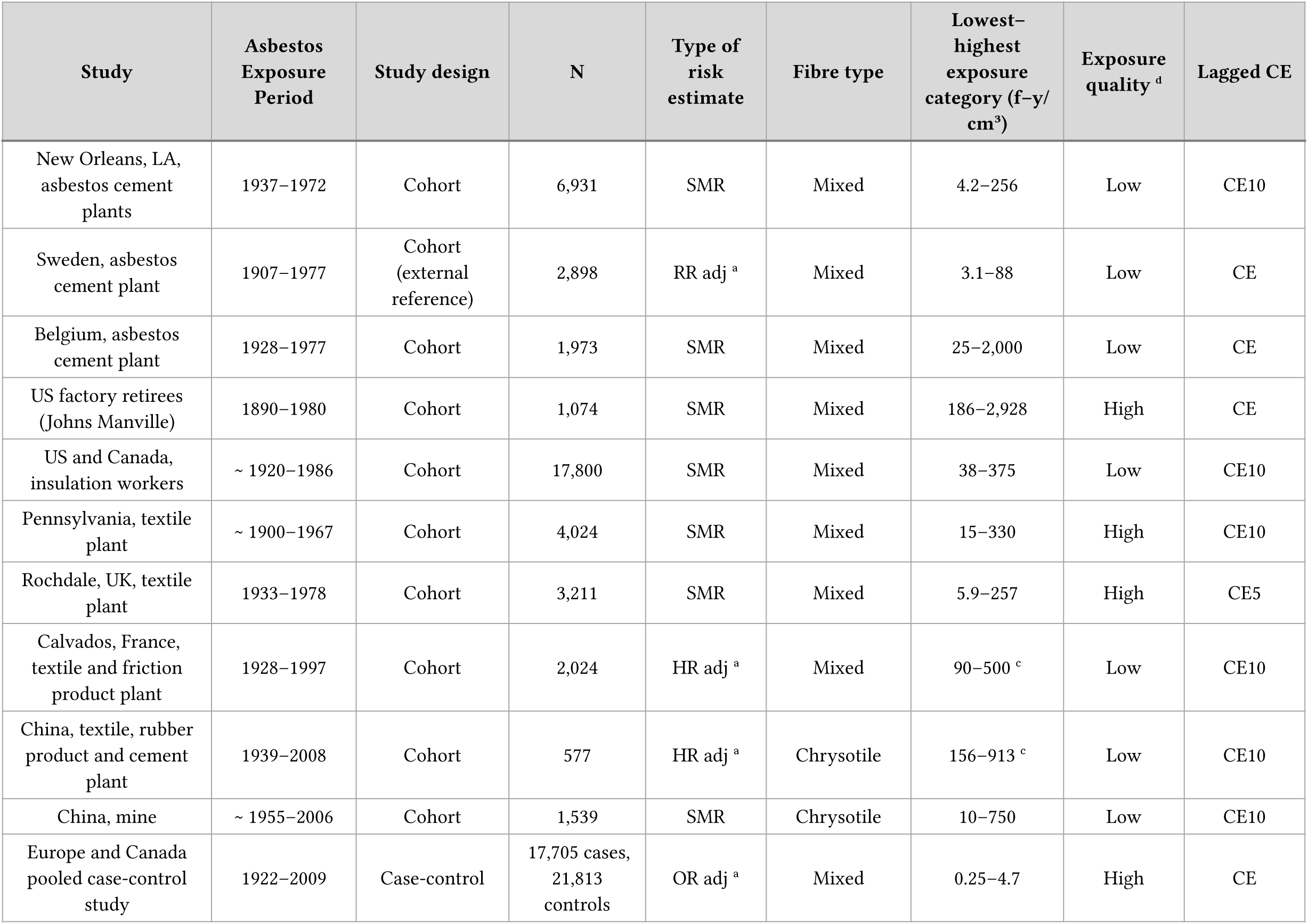
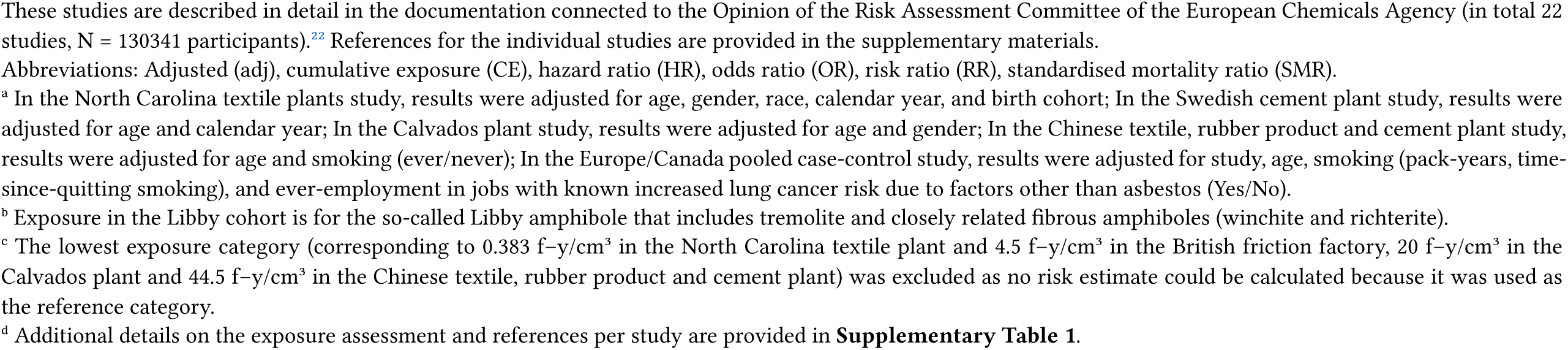
Studies included in the study base for occupational standard setting in Europe by the European Chemicals Agency (ECHA) on quantitative asbestos exposure and lung cancer.

| Study | Asbestos Exposure Period | Study design | N | Type of risk estimate | Fibre type | Lowest–highest exposure category (f–y/cm<sup>3</sup>) | Exposure quality<sup>d</sup> | Lagged CE |
| --- | --- | --- | --- | --- | --- | --- | --- | --- |
| Quebec, Canada, mines and mills | ~ 1904–1976 | Cohort | ~11,000 | SMR | Chrysotile | 4.7–4,710 | Low | CE to age 55 |
| Italy, Balangero, mine and mill | 1916–1990 | Cohort | 1,056 | SMR | Chrysotile | 50–667 | Low | CE |
| Connecticut, friction product plant | 1913–1977 | Cohort | 3,513 | SMR | Chrysotile | 15–400 | Low | CE |
| South Carolina, textile plant | 1896–1977 | Cohort | 3,072 | SMR | Chrysotile | 0.75–200 | High | CE10 |
| North Carolina, textile plant | 1925–1994 | Cohort | 5,770 | RR adj<sup>a</sup> | Chrysotile | 5.7–408<sup>c</sup> | Low | CE10 |
| Wittenoom, Australia, mine | 1937–1966 | Cohort | 6,358 | SMR | Amphibole, crocidolite | 0.11–220 | Low | CE |
| Patterson, NJ, insulation manufacture | 1941–1954 | Cohort | 820 | SMR | Amphibole, amosite | 3–417 | Low | CE |
| Tyler, TX, insulation manufacture | 1954–1972 | Cohort | 1,121 | SMR | Amphibole, amosite | 11–375 | High | CE |
| Libby, MT, mines and mills | 1935–1990 | Cohort | 1,862 | RR | Amphibole, tremolite<sup>b</sup> | 0.7–73 | High | CE20 |
| UK, friction products factory | 1910–1979 | Nested case-control | 13,460 | OR | Mixed | 29.5–228<sup>c</sup> | Low | CE |
| Ontario, asbestos cement plant | 1948–1977 | Cohort | 740 | SMR | Mixed | 15–250 | Low | CE |
| New Orleans, LA, asbestos cement plants | 1937–1972 | Cohort | 6,931 | SMR | Mixed | 4.2–256 | Low | CE10 |
| Sweden, asbestos cement plant | 1907–1977 | Cohort (external reference) | 2,898 | RR adj <sup>a</sup> | Mixed | 3.1–88 | Low | CE |
| Belgium, asbestos cement plant | 1928–1977 | Cohort | 1,973 | SMR | Mixed | 25–2,000 | Low | CE |
| US factory retirees (Johns Manville) | 1890–1980 | Cohort | 1,074 | SMR | Mixed | 186–2,928 | High | CE |
| US and Canada, insulation workers | ~ 1920–1986 | Cohort | 17,800 | SMR | Mixed | 38–375 | Low | CE10 |
| Pennsylvania, textile plant | ~ 1900–1967 | Cohort | 4,024 | SMR | Mixed | 15–330 | High | CE10 |
| Rochdale, UK, textile plant | 1933–1978 | Cohort | 3,211 | SMR | Mixed | 5.9–257 | High | CE5 |
| Calvados, France, textile and friction product plant | 1928–1997 | Cohort | 2,024 | HR adj <sup>a</sup> | Mixed | 90–500 <sup>c</sup> | Low | CE10 |
| China, textile, rubber product and cement plant | 1939–2008 | Cohort | 577 | HR adj <sup>a</sup> | Chrysotile | 156–913 <sup>c</sup> | Low | CE10 |
| China, mine | ~ 1955–2006 | Cohort | 1,539 | SMR | Chrysotile | 10–750 | Low | CE10 |
| Europe and Canada pooled case-control study | 1922–2009 | Case-control | 17,705 cases, 21,813 controls | OR adj <sup>a</sup> | Mixed | 0.25–4.7 | High | CE |

These studies are described in detail in the documentation connected to the Opinion of the Risk Assessment Committee of the European Chemicals Agency (in total 22 studies, N = 130341 participants).<sup>22</sup> References for the individual studies are provided in the supplementary materials.
Abbreviations: Adjusted (adj), cumulative exposure (CE), hazard ratio (HR), odds ratio (OR), risk ratio (RR), standardised mortality ratio (SMR).
<sup>a</sup> In the North Carolina textile plants study, results were adjusted for age, gender, race, calendar year, and birth cohort; In the Swedish cement plant study, results were adjusted for age and calendar year; In the Calvados plant study, results were adjusted for age and gender; In the Chinese textile, rubber product and cement plant study, results were adjusted for age and smoking (ever/never); In the Europe/Canada pooled case-control study, results were adjusted for study, age, smoking (pack-years, time-since-quitting smoking), and ever-employment in jobs with known increased lung cancer risk due to factors other than asbestos (Yes/No).
<sup>b</sup> Exposure in the Libby cohort is for the so-called Libby amphibole that includes tremolite and closely related fibrous amphiboles (winchite and richterite).
<sup>c</sup> The lowest exposure category (corresponding to 0.383 f-y/cm<sup>3</sup> in the North Carolina textile plant and 4.5 f-y/cm<sup>3</sup> in the British friction factory, 20 f-y/cm<sup>3</sup> in the Calvados plant and 44.5 f-y/cm<sup>3</sup> in the Chinese textile, rubber product and cement plant) was excluded as no risk estimate could be calculated because it was used as the reference category.
<sup>d</sup> Additional details on the exposure assessment and references per study are provided in **Supplementary Table 1**.

#### SYNERGY study

SYNERGY is a pooled case-control study on occupational carcinogen risk for lung cancer development, collecting data from 14 case-control studies conducted across Europe and Canada (1985– 2010), comprising 17,705 lung cancer cases and 21,813 controls (**Table 2**). Lung cancer cases with histological or cytological confirmation were included with overall response rate of 83% (range: 62–98%). Controls were mostly recruited from the general population (79% vs 21% hospitals), with a response rate of 70% (range: 41–100%). **Supplementary Table 2** describes sampling schemes for the case-control studies: most were frequency-matched on age and sex from a dynamic population study base, except for MORGEN which is a case-control study nested in the European Prospective Investigation into Cancer and Nutrition study in the Netherlands. It follows that odds ratios (OR) estimated from SYNERGY approximate incidence rate ratios (IRR) under assumptions described and tested in the **Supplementary Methods**.^30^ While IRR may approximate RR under certain circumstances, the use of IRR to estimate PoC could generally overestimate PoC for studies or subpopulations (e.g., older ages) for which outcome is not rare (see **Supplementary Methods**).

**Table 2.**
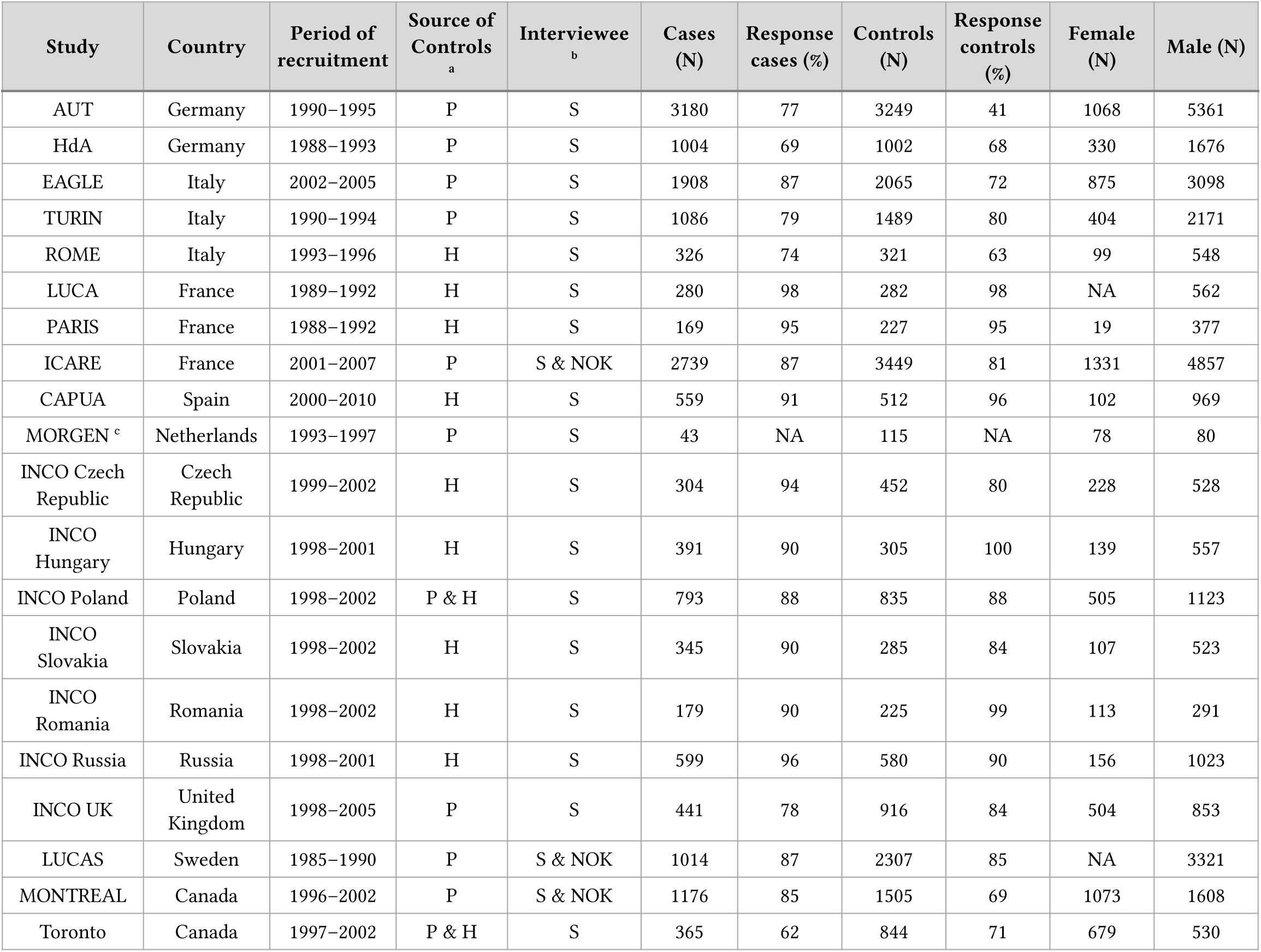
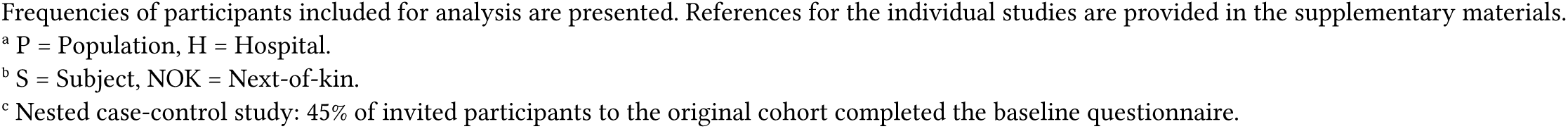
Description of studies included in the SYNERGY project.

| Study | Country | Period of recruitment | Source of Controls<sup>a</sup> | Interviewee<sup>b</sup> | Cases (N) | Response cases (%) | Controls (N) | Response controls (%) | Female (N) | Male (N) |
| --- | --- | --- | --- | --- | --- | --- | --- | --- | --- | --- |
| AUT | Germany | 1990–1995 | P | S | 3180 | 77 | 3249 | 41 | 1068 | 5361 |
| HdA | Germany | 1988–1993 | P | S | 1004 | 69 | 1002 | 68 | 330 | 1676 |
| EAGLE | Italy | 2002–2005 | P | S | 1908 | 87 | 2065 | 72 | 875 | 3098 |
| TURIN | Italy | 1990–1994 | P | S | 1086 | 79 | 1489 | 80 | 404 | 2171 |
| ROME | Italy | 1993–1996 | H | S | 326 | 74 | 321 | 63 | 99 | 548 |
| LUCA | France | 1989–1992 | H | S | 280 | 98 | 282 | 98 | NA | 562 |
| PARIS | France | 1988–1992 | H | S | 169 | 95 | 227 | 95 | 19 | 377 |
| ICARE | France | 2001–2007 | P | S & NOK | 2739 | 87 | 3449 | 81 | 1331 | 4857 |
| CAPUA | Spain | 2000–2010 | H | S | 559 | 91 | 512 | 96 | 102 | 969 |
| MORGEN<sup>c</sup> | Netherlands | 1993–1997 | P | S | 43 | NA | 115 | NA | 78 | 80 |
| INCO Czech Republic | Czech Republic | 1999–2002 | H | S | 304 | 94 | 452 | 80 | 228 | 528 |
| INCO Hungary | Hungary | 1998–2001 | H | S | 391 | 90 | 305 | 100 | 139 | 557 |
| INCO Poland | Poland | 1998–2002 | P & H | S | 793 | 88 | 835 | 88 | 505 | 1123 |
| INCO Slovakia | Slovakia | 1998–2002 | H | S | 345 | 90 | 285 | 84 | 107 | 523 |
| INCO Romania | Romania | 1998–2002 | H | S | 179 | 90 | 225 | 99 | 113 | 291 |
| INCO Russia | Russia | 1998–2001 | H | S | 599 | 96 | 580 | 90 | 156 | 1023 |
| INCO UK | United Kingdom | 1998–2005 | P | S | 441 | 78 | 916 | 84 | 504 | 853 |
| LUCAS | Sweden | 1985–1990 | P | S & NOK | 1014 | 87 | 2307 | 85 | NA | 3321 |
| MONTREAL | Canada | 1996–2002 | P | S & NOK | 1176 | 85 | 1505 | 69 | 1073 | 1608 |
| Toronto | Canada | 1997–2002 | P & H | S | 365 | 62 | 844 | 71 | 679 | 530 |

Frequencies of participants included for analysis are presented. References for the individual studies are provided in the supplementary materials.
<sup>a</sup> P = Population, H = Hospital.
<sup>b</sup> S = Subject, NOK = Next-of-kin.
<sup>c</sup> Nested case-control study: 45% of invited participants to the original cohort completed the baseline questionnaire.

Cumulative occupational asbestos exposure was estimated using the quantitative *SYN-JEM*,^31^ developed using 27,958 asbestos exposure measurements. SYN-JEM estimates annual average job (ISCO-68), year and region-specific asbestos exposure in fibre-year (ff/ml-year) which were consequently summed to yield cumulative exposure in fibre-years. The SYN-JEM model incorporated effects of country-specific asbestos bans and has been applied in studies on occupational asbestos exposure and asbestos-related diseases: peritoneal mesothelioma,^32^ asbestosis,^33^ and lung cancer.^26^

Ethical approvals were obtained according to each country’s legislation and by the International Agency for Research on Cancer (IARC) institutional review board (IEC 07-05). Participants gave informed consent to participate in the study before taking part.

### Statistical Analysis

#### Exposure-response estimation

Exposure-response relations in the 22 occupational asbestos studies were estimated with random-effects meta-regression of study-level logRR and average exposure level per category reported. A meta-RR was estimated from meta-regression of fixed-effects estimates (**Supplementary Methods**).^20^

A one-stage meta-analysis^34^ of SYNERGY was used to estimate the relation between lung cancer RR and lifetime cumulative asbestos (**Supplementary Methods**). Meta-estimates were obtained with a generalised linear mixed model (logit link) with study-specific random intercepts and random slopes for cumulative asbestos exposure in fibre-years to account for between-study heterogeneity. The model was adjusted by including fixed effects for sex, study, age group (<45, 45–49, 50–54, 55–59, 60–64, 65–69, 70–74, and >74 years), smoking (cigarette pack-years [log (cigarette pack-years +1)] and time-since-quitting smoking cigarettes (current smokers; stopping smoking 2–7, 8–15, 16–25, ≥26 years before interview/diagnosis; and never-smokers).

#### Heterogeneity and prediction intervals

Between-study heterogeneity was quantified using the I^2^ statistic, representing the proportion of variation attributable to genuine differences between studies rather than sampling error.^35^ In SYNERGY, exposure-response heterogeneity was obtained from two-stage univariate meta-analysis (**Supplementary Methods**). Higher I^2^ values indicate greater inconsistency in estimated effects across studies beyond expected by chance.

For both meta-analyses, 95% prediction intervals (PI) were estimated using the approach described by Higgins, et al.^36^ assuming normally distributed effects. This method combines squared standard error of the target effect with estimated between-study variance. For occupational studies, the target was the pooled logRR and in SYNERGY, the asbestos exposure-response slope coefficient. The prediction interval represents the range where the true effect estimate from a new study would be expected to fall, accounting for both sampling uncertainty and between-study heterogeneity as uncertainty measure.

#### Probability of Causation

PoC for individual cases is the probability of necessity, that is, the probability that the disease would not have occurred without the exposure.^11^ Its lower bound is the excess risk among the exposed divided by their risk, (Risk_exp_ − Riskun_exp_)/Risk_exp_, which re-expressed as a risk ratio is (RR−1)/RR. Both risks are cumulative probabilities of lung cancer over the same defined period. For quantitative exposures, PoC has been estimated using RR at exposure value (*x*) from the exposure-response relation:^7,8^

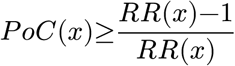

This equation was solved for *P oC*(*x*) ≥ 50% for both exposure-response relations to obtain exposure compensation thresholds. Lung cancer cases with PoC ≥ 50% are reported as total count of compensated cases and rate per 10,000 cases using the SYNERGY population as reference. Besides PoC point estimates, upper limits of 95% PI were estimated to account for uncertainties in exposure-response relations (*presumably plausible* threshold).

#### Sensitivity Analysis

The exposure-response in occupational asbestos studies was re-estimated by excluding SYNERGY. SYNERGY exposure-response relations were re-estimated stratifying by control source (hospital-based and population-based), since hospital-based controls have greater baseline disease risk than population-based controls, resulting in potential effect underestimation. Additional sensitivity analyses considered alternative meta-analysis configurations and effect modifiers of the exposure-response (**Supplementary Methods**).

Analyses and visualisations were performed in R v.4.5.1 through RStudio v.2025.05.1. Code and documentation are available at https://github.com/UtrechtUniversity/PoC-Asbestos.

## Results

Most occupational studies (in total 22 studies, N = 130341 participants) are cohorts estimating standardised mortality ratios for a range of exposure categories (**Table 1**). Asbestos exposure periods range from 1890 to 2009 (17 studies with exposure starting before World War II; **Supplementary Figure 2**). Risk estimates correspond to cumulative asbestos exposures up to 4710 fibre-years (median 79.9, IQR 17.5 – 250.3; excluding SYNERGY: median 90, IQR 25.5 – 256.3). Exposure assessment quality was rated high in 7 studies and low in 15 due to poor documentation, low exposure contrast, use of external instead of study-specific fibre conversion factors, incomplete job histories, or incomplete exposure coverage during follow-up.21

A summary of the fourteen studies included in SYNERGY is shown in **Table 2**, and characteristics of the study population (N = 37866) in **Table 3**. Exposure to asbestos for the SYNERGY participants had to be estimated from 1922 to 2009. Ever exposure to asbestos was reported in 39% (n = 14752) participants. Cumulative asbestos exposure in SYNERGY ranged from 0.002 to 64.6 fibre-years (median 1.33, IQR: 0.55 – 3.22). The percentage of asbestos exposure in lung cancer cases and controls was 44% (n = 7440/16901) and 34.9% (n = 7312/20965), respectively. Median lifetime cumulative exposure to asbestos in ever-exposed was 1.54 (IQR: 0.62 – 3.53, range: 0.002 – 35.43) fibre-years in cases and 1.17 (IQR: 0.48 – 2.86, range: 0.002 – 64.6) fibre-years in controls. Total exposure duration in the cases and controls was 15.5 (IQR: 6 – 31, range: 1 – 62) years and 14 (IQR: 5 – 30, range: 1 – 63) years, respectively.

**Table 3.** Descriptive characteristics of SYNERGY study participants.

|  | Total (N=37866) | Controls (N=20965) | Lung cancer cases (N=16901) |
| --- | --- | --- | --- |
| **Sex** |  |  |  |
| Female | 7810 (20.6%) | 4514 (21.5%) | 3296 (19.5%) |
| Male | 30056 (79.4%) | 16451 (78.5%) | 13605 (80.5%) |
| **Age** |  |  |  |
| Mean (SD) | 61.7 (9.63) | 61.5 (9.92) | 62.0 (9.23) |
| **Asbestos (ff/ml-years)** |  |  |  |
| Mean (SD) <sup>a</sup> | 2.42 (3.01) | 2.22 (3.01) | 2.60 (3.00) |
| Median (Q1, Q3) <sup>a</sup> | 1.33 (0.550, 3.22) | 1.17 (0.476, 2.86) | 1.54 (0.622, 3.53) |
| Min, Max <sup>a</sup> | 0.002, 64.6 | 0.002, 64.6 | 0.002, 35.4 |
| Never exposed | 23114 (61.0%) | 13653 (65.1%) | 9461 (56.0%) |
| **Exposure duration (years)** |  |  |  |
| Mean (SD) <sup>a</sup> | 18.4 (13.9) | 17.9 (13.8) | 18.9 (14.0) |
| Median (Q1, Q3) <sup>a</sup> | 15.0 (6.00, 30.0) | 14.0 (5.00, 30.0) | 15.5 (6.00, 31.0) |
| Min, Max <sup>a</sup> | 1.00, 63.0 | 1.00, 63.0 | 1.00, 62.0 |
| Never exposed | 23114 (61.0%) | 13653 (65.1%) | 9461 (56.0%) |
| **Smoking** |  |  |  |
| Never smoker | 8522 (22.5%) | 7153 (34.1%) | 1369 (8.1%) |
| Former smoker | 13652 (36.1%) | 8220 (39.2%) | 5432 (32.1%) |
| Current smoker | 15692 (41.4%) | 5592 (26.7%) | 10100 (59.8%) |
| **Pack-years** |  |  |  |
| Median (Q1, Q3) | 23.3 (1.60, 41.5) | 9.75 (0, 29.1) | 35.8 (21.0, 51.0) |
| **Time since quitting smoking** |  |  |  |
| 0-7 years | 3448 (9.1%) | 1422 (6.8%) | 2026 (12.0%) |
| 8-15 years | 3429 (9.1%) | 1898 (9.1%) | 1531 (9.1%) |
| 16-25 years | 3517 (9.3%) | 2346 (11.2%) | 1171 (6.9%) |
| >25 years | 3258 (8.6%) | 2554 (12.2%) | 704 (4.2%) |

<sup>a</sup> Calculated among the ever-exposed.
Abbreviations: fibre-years (ff/ml-years), maximum (Max), minimum (Min), 25th percentile (Q1), 75th percentile (Q3), standard deviation (SD).

Cumulative exposure in studies with hospital-based controls had comparable exposure on average in cases and controls (median 2.39 fibre-years in cases and 2.27 in controls) while in studies with population-based controls the cases had a higher median cumulative exposure than controls (1.31 vs 0.98 fibre-years, **Supplementary Table 3**). The difference in smoking intensity among ever-smokers was greater in hospital-based control studies (37.8 pack-years in cases vs 27.0 in controls) than population-based control studies (37.9 vs 21.6 pack-years, **Supplementary Table 4**).

### Exposure-response and PoC

In the meta-analysis of the 22 occupational studies (I^2^ = 92.7%), lung cancer RR doubled at 62.9 fibre-years, and 18.2 fibre-years when using the upper limit of the 95% PI (**Figure 1A**). If these thresholds were to be applied to the lung cancer cases and their cumulative asbestos exposure estimate from the SYNERGY population, 0 and 17 lung cancer cases per 10,000 would be entitled to compensation under the point estimate and presumably plausible threshold, respectively (**Table 4**). Exclusion of SYNERGY from the database yielded an exposure-response relation with similar heterogeneity (I^2^ = 92.9%) and lung cancer risk doubling threshold of 84.1 fibre-years and 26.6 with the upper 95% PI (**Supplementary Table 5**).

**Figure 1.**
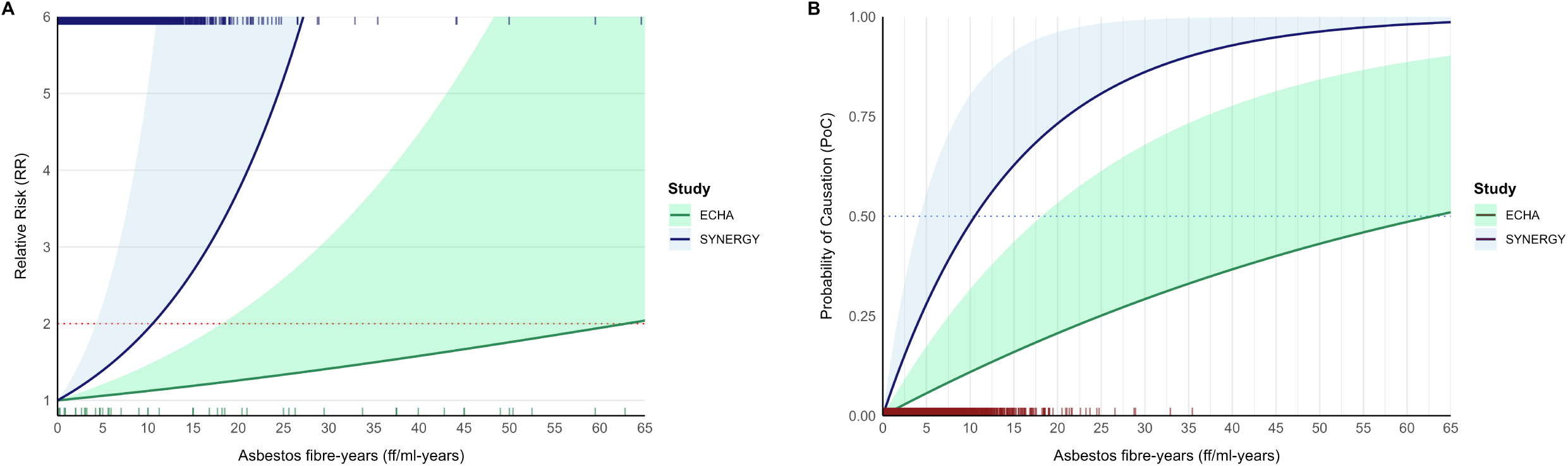
Exposure response relations of lung cancer risk increase (**A**), and Probability of Causation of lung cancer (**B**) across the range of lifetime cumulative exposure to asbestos (fibre-years) values in SYNERGY. Solid lines represent the average meta-estimates; shaded areas, the upper 95% prediction intervals, and the dotted lines, the doubling in increase of risk threshold (red) and PoC ≥ 50% threshold (blue). Vertical tick marks represent average exposure values for the reported pooled estimates per exposure category in the study base by ECHA (green, bottom), individual participant exposure values in the total SYNERGY population (blue, top), and lung cancer cases in SYNERGY (red, bottom).

**Table 4.** Number of compensated cases per 10,000 lung cancer cases according to various scenarios applied to the lung cancer cases (n = 16901) within the SYNERGY population. It was estimated at what cumulative asbestos exposure (fibre-years) the probability of causation (PoC) is equal or larger than 50%.

| Model<sup>a</sup> | Risk Increase per Fibre-year<sup>b</sup> |  | Min Exposure for 50% PoC (fibre-years)<sup>c</sup> |  | Cases per 10,000 above 50% PoC in SYNERGY |  |
| --- | --- | --- | --- | --- | --- | --- |
|  | Estimate | 95% Prediction Interval | Point Estimate | Presumably Plausible | Point Estimate | Presumably Plausible |
| **Meta-regression of published occupational studies including SYNERGY** |  |  |  |  |  |  |
| Spline | 1.1% | [0%, 3.6%] | 62.93 | 18.19 | 0 | 17 |
| **Meta-analysis of SYNERGY studies only** |  |  |  |  |  |  |
| Main | 6.8% | [0%, 17.5%] | 10.52 | 4.30 | 117 | 846 |
| **Sensitivity analysis of SYNERGY studies, stratified by source of controls** |  |  |  |  |  |  |
| Population controls | 10.2% | [3.7%, 17.0%] | 7.16 | 4.41 | 344 | 820 |
| Hospital controls | 2.3% | [0%, 8.6%] | 29.92 | 8.40 | 1 | 223 |

<sup>a</sup> **Spline**: random-effects meta-regression of the 22 occupational studies, with a natural spline for cumulative exposure (Supplementary Methods, Equation 1). **Main**: one-stage meta-analysis of SYNERGY with random study intercepts and random slopes for cumulative exposure, linear in fibre-years (Supplementary Methods, Equation 3).
<sup>b</sup> Negative values truncated to 0 due to monotonicity assumption. For spline model: instantaneous slope at 1.5 fibre-years (median exposure in cases).
<sup>c</sup> Probability of Causation (PoC) of 50% corresponds to a doubling in lung cancer risk (i.e., RR = 2) for the point estimate and the upper 95% prediction interval for the presumably plausible threshold.

The exposure-response relation in SYNERGY (I^2^ = 63.4%) modelled as a linear function of cumulative asbestos exposure (fibre-years) yielded a 6.8% lung cancer risk increase per fibre-year, with upper PI of 17.5% (**Figure 1A**). Study-specific estimates and results of the two-stage meta-analysis are presented in **Supplementary Figure 3** and **Supplementary Table 6**. In the one-stage meta-analysis, lung cancer risk is doubled at 10.5 fibre-years for the point estimate and 4.3 fibre-years under the presumably plausible threshold (**Figure 1B**), resulting in 117 and 846 compensated cases per 10,000, respectively (**Table 4**).

In the sensitivity analysis, a steeper exposure-response (**Supplementary Figure 4**) was observed for studies with population-based controls (10.2% risk increase per fibre-year) compared to studies with hospital controls (2.3%), with large shifts in PoC ≥50% thresholds (**Supplementary Figure 5**) for the point estimates, but limited impact in the presumably plausible threshold for population controls compared to the average SYNERGY estimate (**Table 4**). Of ten candidate modifiers, only source of controls (steeper in population-based controls) and smoking status (steeper in current smokers) modified the exposure-response. Presumably plausible thresholds ranged from 3.4 to 5.9 fibre-years across strata of all modifiers other than source of controls (**Supplementary Tables 7–9**).

## Discussion

In this study, we analysed two large datasets to provide a transparent and reproducible PoC estimate of asbestos-related lung cancer, as required by the Dutch compensation scheme, to determine exposure thresholds for financial compensation of lung cancer cases occupationally exposed to asbestos. We used occupational asbestos studies, mainly industrial cohorts, which form the core evidence on quantitative occupational asbestos exposure and (lung) cancer risk, and a more recently conducted, pooled general population case-control study. Contrary to expectations given the traditional view of cohort studies as providing stronger evidence, the case-control studies proved more informative for determining compensation thresholds. The cohort studies are representative of high asbestos exposure (median 90 fibre-years) in production (mines and factories) industries, compared to the case-control studies in SYNERGY exposed to significantly lower levels of mixed asbestos fibres (median 1.3 fibre-years) in industries using asbestos products (e.g., construction, maintenance). Meta-analysis of industrial cohort studies showed a relatively shallow exposure-response relation, clearly levelling off at higher exposures leading to a higher exposure threshold for PoC ≥50% than the Helsinki criteria which assumed a linear increasing risk with increasing exposure. Despite higher heterogeneity among occupational cohort studies, the combination of shallow exposure response and higher heterogeneity led to higher, not lower, exposure thresholds when prediction intervals were used, compared to the general population pooled case-control studies. SYNERGY showed a considerably steeper exposure-response relation, resulting in lower thresholds, particularly when using upper prediction intervals. Several aspects determine which estimates should be used for compensation:

- **Study quality, including the exposure assessment component**. An important limitation of the occupational cohort studies meta-regression is the clear indication of exposure-response underestimation due to individual study limitations, particularly varying exposure assessment quality. Differences in study quality among the occupational cohort studies likely explain much of the between-study heterogeneity (I^2^ = 92.7%).^21^ This issue is widely recognised in occupational epidemiology due to exposure misclassification and measurement error, discussed in the context of asbestos.^21,37^ Other potential biases include confounding from unmeasured confounders, particularly cigarette smoking, and possibly selection bias due to the healthy worker effect in the occupational cohort studies, the latter often resulting in effect underestimation.^38^ Healthy-worker survivor bias has been shown in one of the included cohorts.^39^ Asbestos also increases the risk of competing causes of death, such as mesothelioma. Therefore, both the occupational cohort studies and SYNERGY estimate the effect of asbestos on lung cancer among those who did not die from competing causes, rather than its total effect on lung cancer risk.^40^ In SYNERGY, this may be accentuated because only three studies allowed next-of-kin responses for deceased cases. Matching on age limits age-related differences in survival between cases and controls, but not the exposure-related depletion of participants due to competing causes of death.
- Cumulative asbestos exposure in SYNERGY is based on SYN-JEM,^31^ which has the advantage of assigning values with one method based on modelled personal measurements of asbestos exposure across the pooled case-control studies. However, it potentially introduces Berkson-type error by using average exposure values,^41^ resulting in imprecise (wider confidence intervals) exposure-response estimation.^42^ Berkson-type error is also present in industrial cohort studies since exposures are also assigned at job level, but often at a more refined level than in general population studies (company-specific job codes versus ISCO-68). Unlike industrial cohort studies, in which each study used its own exposure assessment method, SYNERGY used the same job-exposure matrix (SYN-JEM) across centres, estimating exposure levels specific to job, calendar year and region, showing less effect heterogeneity (I^2^ = 63.4%). While most measurements used to develop SYN-JEM used phase contrast microscopy (PCM) with a lower limit of detection (LOD) of ∼2,000 fibres/m^3^, 5% used electron microscopy (EM) with LOD 100 to 200 fibres/m^3^.^23^ In contrast, only one^28^ of the industrial cohort studies used EM besides PCM and particle counting, whereas many applied conversion factors due to changes in analytic techniques over time, some study specific, some assessed in other studies.^21^
- **Other reasons for study heterogeneity**. Exposure to different asbestos types may have contributed to between-study differences. However, potency differences between fibre types are considerably smaller for lung cancer than for mesothelioma.^43^ Moreover, the analysis by van der Bij et al.^20^ using a spline model instead of assuming linearity of the exposure response relation, as well as the analysis by ECHA,^22^ suggest that the potency differences for lung cancer are even smaller than previously indicated by simple linear models rather than more advanced spline models, which more adequately describe the exposure-response relation. In SYNERGY, the exposure-response was steeper among current smokers. This was not used to set compensation thresholds, as it would compensate smokers at lower asbestos exposures than never-smokers with the same occupational exposure.^7^ Future research could apply causally interpretable meta-analysis methods to transport exposure-response estimates to a Dutch target population,^44^ which would require individual participant data and covariate data from the Dutch working population.
- **Relevance for asbestos risk assessment**. The industrial cohorts describe lung cancer risk at the high end of the exposure distribution, whereas case-control studies describe risk at considerably lower exposure levels. Most industrial cohorts included workers who started working before World War II and have been updated with additional follow-up, resulting in higher numbers of deceased workers and increased power to detect associations. This allowed more detailed exploration of the shape of the association between exposure and disease. However, exposure assessment for these early periods is inherently uncertain, as systematic personal air sampling was not routinely conducted before the 1970s. Nonetheless, industrial cohort studies often excluded workers with short employment duration (i.e., <1 year employed). The number of observations at the low end of the exposure distribution (e.g., below 25-50 fibre-years) is limited for the occupational cohorts and knowledge about the risk at this level is strongly dependent on extrapolation to lower exposure levels and models used for extrapolations, which mostly assumed linearity. Moreover, most exposure in the occupational cohorts occurred more than half a century ago (**Supplementary Figure 2**), in populations with a shorter life expectancy and different mortality patterns, including competing causes of death, than the more recently recruited SYNERGY populations. Besides exposure assessment quality, this may also contribute to differences in exposure-response between both study bases.
- Initial observations of increased asbestos health risk emerged in mining and industries processing asbestos in products like textile, asbestos cement, brake lining, and shipyards (asbestos insulation). Currently, elevated risk occurs in workers from industries not involved in primary asbestos production. A recent UK mesothelioma case-control study shows a clear shift in risk for the working population from primary production, where the first excess risks were observed, due to occupational asbestos exposure, to downstream industries, such as construction, since World War II.^45^

On the basis of these analyses, the Advisory Committee for the List of Occupational Diseases in the Netherlands advised the Minister of Social Affairs and Employment that the lifetime cumulative exposure threshold of 4.3 fibre-years based on SYNERGY meta-estimates to compensate workers occupationally exposed to asbestos is defensible. This threshold aligns better with the expected burden of asbestos-related lung cancer in the Netherlands. The most recent general population estimate projects 600 new asbestos-related lung cancer cases, based on registered mesothelioma cases and an assumed lung cancer/mesothelioma ratio,^17^ out of 14,089 lung cancer cases diagnosed nationally in 2023,^46^ or approximately 426 per 10,000 cases. Applied to the cumulative exposure of the SYNERGY lung cancer cases, the 4.3 fibre-year threshold would compensate 846 per 10,000 cases (117 per 10,000 at the point estimate), whereas the thresholds from occupational cohort studies would compensate 0 to 17 per 10,000, far below this expected burden. As a secondary consideration, Dutch workers currently applying for compensation were more recently exposed at lower mixed asbestos fibre levels.

The PoC is easily communicated to medical specialists, occupational health specialists, patients, and stakeholders as an eligibility criterion for financial compensation. Nonetheless, harm definition is linked to a doubling in lung cancer risk.^47^ Asbestos is a no-threshold lung carcinogen, meaning any exposure level can increase lung cancer risk. Therefore, PoC ≥50% should not be interpreted as definitive evidence that lung cancer was caused by asbestos, but as a commonly used public health decision-making tool. While exposure-response-based PoC allows decisions with individual cumulative exposure, it relies on population-average effects. Because exposure may accelerate disease onset and background risk varies between individuals, (RR−1)/RR can underestimate the true PoC.^9,10^ Conversely, using rate rather than risk ratios, without a defined risk period, may overestimate PoC where lung cancer is common, such as at older ages, where the exposure-response also appeared shallower (**Supplementary Table 9**). Applying uncertainties in the favour of claimants for workers’ compensation has been common practice to balance the limitations of the PoC.^4,6,7^ A single PoC threshold may however overcompensate some subgroups, such as older adults, and undercompensate others, although lower thresholds in some subgroups may partly reflect wider prediction intervals due to smaller sample sizes. We discuss the assumptions underlying these estimates with strengths and limitations in greater detail in the **Supplementary Materials**. Future directions for legal applications and workers’ compensation could involve decision-making using health outcomes accounting for accelerated occurrence^48^ and different dimensions of affection, such as loss of life expectancy,^49^ years of working life lost,^50^ disability-adjusted life years,^51^ or loss of income.^52^

## Conclusions

SYNERGY meta-analysis likely provided more relevant estimates of asbestos-related lung cancer for the Dutch compensation scheme than meta-regression of industrial cohort studies, given that current occupational exposure in countries such as the Netherlands involves lower levels of generally mixed asbestos fibres. Steeper exposure-response relation in SYNERGY with lower between-study heterogeneity translates into lower cut-off for compensation using the upper bound of the 95% prediction interval of PoC. Consequently, an exposure threshold of 4.3 fibre-years was selected for the one-time compensation of workers occupationally exposed to asbestos in the Netherlands who develop lung cancer.

## Data Availability

Data are available upon reasonable request. Inquiries can be directed to Dr. Susan Peters (s.peters[at]uu.nl). Alternatively, https://synergy.iarc.who.int/contact/. The source code for this manuscript, statistical analyses, and project documentation are available through the online repository: https://github.com/UtrechtUniversity/PoC-Asbestos.

## Acknowledgements

We extend our sincere appreciation to the SYNERGY study group and the principal investigators of the original studies: https://synergy.iarc.fr/collaborators/.

## Ethics statement

The SYNERGY study was approved by the IARC Ethics Committee (IEC 07-05) in addition to the individual study ethics approvals. Participants gave informed consent before taking part.

## Data and code availability

Data are available upon reasonable request (enquiries can be directed to Dr. Susan Peters. Alternatively, https://synergy.iarc.who.int/contact/.

The source code for this manuscript, statistical analyses, and project documentation is available through the online repository: https://github.com/UtrechtUniversity/PoC-Asbestos.

## AI-use statement

Claude (Anthropic) was used to assist with rephrasing text and improving English grammar and style, and in adapting and documenting analysis code. All AI-generated content was reviewed and edited by the authors, who take full responsibility for the final content

