## Supplementary Materials for "Probability of causation in individual workers: Lung cancer due to occupational exposure to asbestos"

Javier Mancilla-Galindo[1], Susan Peters[1], Huifang Deng[1], Henk van der Molen[2], Hans Kromhout[1], Lützen Portengen[1], Roel Vermeulen[1], Dick Heederik[1,3]

1. Institute for Risk Assessment Sciences, Utrecht University, Utrecht, the Netherlands
2. Department of Public and Occupational Health, Amsterdam UMC, Amsterdam, the Netherlands
3. National Expertise Centre for Substance-related Occupational Diseases (Lexces), Utrecht, the Netherlands

### Supplementary Methods

#### Causal Model

A causal model of the effect of cumulative exposure to asbestos (fibre-years) on lung cancer risk was represented in a directed acyclic graph (DAG) (**Supplementary Figure 1**) and deposited in OpenCausal.org.[1] Asbestos exposure accumulates as the product of intensity and duration, and in a causal framework it decomposes into a series of time-varying point exposures of different magnitudes (not shown in this DAG for simplicity). Under that decomposition, employment status is simultaneously a mediator of earlier exposure and a confounder of later exposure, which opens the healthy-worker survivor bias. Naimi, et al.[2] assessed the three components of this bias in the South Carolina chrysotile textile cohort, and found all three present; neither exposure lagging nor adjustment for employment status removed it. Co-exposure to other occupational carcinogens is represented with RCS as a motivating example. True exposure is unmeasured and estimated with SYN-JEM, resulting in loss of individual variation (Epsilon), and exposure error terms (U) that are independent of outcome status (Berkson-type error).[3] Age, sex, smoking status and intensity, and time since quitting act as confounders. Study centre is considered a proxy for other study-level covariates. Occupational lung carcinogens are assumed to be associated through the common cause industry which serves as a proxy for shared working environment conditions. This backdoor pathway was intentionally not closed as co-exposure to occupational lung carcinogens is frequent.[4] Estimating conditional effects by setting such co-exposures to zero could violate positivity (if certain covariate combinations never occur for a given job title) and consistency (counterfactual scenarios such as high asbestos exposure with zero RCS are implausible in some industries). Monotonicity is assumed throughout this analysis, implying that asbestos cannot prevent lung cancer through any causal mechanism, as well as no-interference (only individual exposure is relevant for the effect and not affected by others' exposures). Lastly, both age and sex are matching factors potentially causing selection bias (not shown in this DAG for simplicity). Adjustment for these matching variables through unconditional logistic regression results in unbiased estimates under the rare disease assumption.[5]

#### Exposure-response estimation

For the 22 occupational asbestos studies, four different combinations of modelling assumptions were examined (linear and natural splines, allowing for differences or no differences in background lung cancer risk) as described in van der Bij, et al.[6] Exposure-response meta-regression models were fitted using restricted maximum likelihood (REML) estimation with random effects to account for between-study heterogeneity. Study-specific log-transformed relative risks (logRR) were modelled as a function of the mean exposure level within each reported category. Relative risk estimates with 95% confidence and prediction intervals were generated for exposure levels ranging from 0 to 200 fibre-years to compare the four models (**Supplementary Figure 6**). The spline meta-regression model with fixed intercept was used for further analysis due to reasons explained elsewhere,[6] for the estimation of $RR(x)$ using the natural spline (NS) basis functions estimated using third-order polynomials (Equation 1).

$$RR(x) = \exp(\beta_1 \cdot NS_1(x) + \beta_2 \cdot NS_2(x)) \quad (1)$$

For the SYNERGY studies, two different individual participant data (IPD) meta-analyses were conducted.[7] Motivation for each meta-analysis is given below alongside its description.

**1. Two-stage univariate meta-analysis**

The two-stage univariate random-effects meta-analysis estimates the effect of exposure on outcome per study with IPD in the first stage, and the resulting pooled effects are synthesised in the second stage. This process is equivalent to a traditional random-effects meta-analysis, ignoring within-study and between-study covariance in parameter estimates, which can be problematic and lead to bias.[7] Conventional measures of heterogeneity ($I^2$) can be estimated with this approach and most readers are familiar with the presentation of this type of meta-analysis.

In the first stage, each study was analysed separately by fitting unconditional multivariable logistic regression models of the effect of cumulative exposure to asbestos (fibre-years) on lung cancer risk (relative risk per fibre-year), adjusted for confounders (Equation 2). Study-specific maximum likelihood models accounted for differences in eligibility criteria and the available contrast in confounders per study, as two included men only (LUCA[8] and LUCAS[9]), one oversampled never-smokers (Toronto[10]), and one collected smoker and never-smoker status but not time since quitting smoking (MORGEN[11]).

$$y_i \sim \text{Bernoulli } (p_i)$$

$$\text{logit } (p_i) = \alpha + \beta x_i + \sum_{k=1}^{K} \theta_k z_{ik} \quad (2)$$

where:

- $y_i$ = Lung cancer status for individual $i$
- $x_i$ = Cumulative asbestos exposure in fibre-years (primary exposure of interest) for individual $i$
- $z_{i1}$ = Age group (always included as categories <45, 45–49, 50–54, 55–59, 60–64, 65–69, 70–74, and >74 years)
- $z_{i2}$ = Sex (included if both men and women included in the study)
- $z_{i3}$ = Cigarette pack-years (always included, as [log (cigarette pack-years +1)])
- $z_{i4}$ = Time since quitting smoking cigarettes (included if more than one category contrast present: current smokers; stopping smoking 2–7, 8–15, 16–25, ≥26 years before interview/diagnosis; and never-smokers)
- $\alpha$ = intercept
- $\beta$ = log odds ratio per asbestos fibre-year (parameter of primary interest)
- $\theta_k$ = log odds ratios for covariates
- $k$ varies from 2 to 4 depending on the available covariates with contrasting levels per study.

Fully adjusted models were obtained for 18 out of the 20 substudies, whereas two studies were not adjusted for sex due to the lack of contrast (LUCA[8] and LUCAS[9]). For the second stage, we fitted REML random-effects meta-analysis of the pooled effect estimates obtained in the first stage. Study-specific estimates and pooled measures of effect are presented in a forest plot (**Supplementary Figure 3**).

#### 2. One-stage meta-analysis

One-stage meta-analysis allows for the simultaneous analysis of all IPD with a mixed-effects model in a flexible framework, which can improve precision of the estimates. We attempted to fit the extended one-stage models described by Debray, et al.[7] using mixed-effects logistic regression models with adaptive Gauss-Hermite Quadrature with 1 quadrature point (Laplacian approximation), including cumulative asbestos as the main explanatory variable, accounting for clustering within each study, allowing for random slopes for cumulative asbestos within study in all model specifications, and adjusting for age group, sex, cigarette pack-years, and time since quitting smoking. A fully-random effects model where all parameters (including intercepts) have random effects with potential correlations, and a reduced random-effects model with independent random effects for all variables failed to converge. A simplified model with random slopes per study for the main variable of interest (cumulative asbestos), random intercepts per study, and fixed effects for all other covariates is thus presented as the main meta-analysis.

For frequency-matched case-control studies, intercepts are not expected to represent baseline disease risk, and are largely reflective of sampling ratios per study. However, differential participation rates across studies can also alter the intercepts, reason why intercepts cannot be considered completely uninformative. Currently, there is not definitive guidance on the best way to analyse case-control IPD regarding the impact of assuming random or stratified intercepts. Nonetheless, guidance for IPD meta-analysis of randomised controlled trials suggests that non-parametric modelling of intercepts avoids potentially strong assumptions on the distribution of intercepts and their influence on the estimated effect.[12] Therefore, we fitted the same model as described before, but including stratified intercepts per study instead of random intercepts as a sensitivity analysis.

For both models, the $RR(x)$ was obtained from the main cumulative asbestos exposure effect $\beta$ (Equation 3). A comparison of the analysis of SYNERGY data under different meta-analyses and assumptions is presented in **Supplementary Table 6**.

$$RR(x) = \exp(\beta \cdot x) \quad (3)$$

#### Conditions for interpreting the estimated odds ratio

The probability of causation is defined on risks, that is, on the probability that the outcome occurs over a defined period. A clearly defined risk period is not available for most of the SYNERGY pooled case-control studies (see **Supplementary Table 2**). For a case-control analysis that is not stratified on sampling time, Kerr, et al.[13] give three sufficient conditions for interpretation of odds ratios as an estimator of the incidence rate ratio (IRR): the population prevalence of exposure is constant over the study period; the hazard ratio (HR) is constant over that period; and no selection bias. For studies matching on time, Greenland and Thomas showed that the odds ratio is a consistent estimator of the IRR provided only that the HR is approximately constant over the study period and that control selection is independent of exposure.[14]

We assessed stability of exposure prevalence among controls in 18 centres of the SYNERGY pooled case-control study (excluding LUCAS, in which controls were also matched on year of inclusion, and MORGEN, nested in a cohort; **Supplementary Table 2**). The prevalence of ever-exposure to asbestos drifted by a median of 4.6 percentage points (IQR 3–8.2) across each centre's own recruitment window, and the exposure odds ratio per calendar year, pooled across all centres, was 0.979 (95% CI 0.926–1.035), consistent with no systematic drift. Greenland and Thomas show that the bias arising from a violation of this condition is a function of the magnitude of the variation in the proportion exposed, and is negligible unless that variation is very large.[14] We additionally assessed the condition that the control series represent person-time over the period in which the cases arose.[15] Comparing each centre's control interview years against its own case diagnosis window, a median of 93.6% of controls per centre were interviewed inside that window (IQR 78–100; 18 centres). The constancy of the hazard ratio over the recruitment window was assessed as a within-centre interview-year interaction on the asbestos slope, reported with the other effect-modification analyses in **Supplementary Table 7**.

An important consideration is the relation between the IRR and the risk ratio over a specified period. Let $[t_0, t_1]$ denote this period, $R_0(t_0, t_1)$ the risk (cumulative incidence) of lung cancer among the unexposed over this period, and $R_1(t_0, t_1)$ the corresponding risk among the exposed. If the hazard ratio is constant over the period, so that the IRR is the same in every part of it, and competing risks are negligible, then $R_1(t_0, t_1) = 1 - [1 - R_0(t_0, t_1)]^{\text{IRR}}$ and the risk ratio is

$$RR(t_0, t_1) = \frac{R_1(t_0, t_1)}{R_0(t_0, t_1)} = \frac{1 - [1 - R_0(t_0, t_1)]^{\text{IRR}}}{R_0(t_0, t_1)} \quad (4)$$

When lung cancer is rare over the period, $RR(t_0, t_1)$ is approximately equal to the IRR.[16] Otherwise, for a harmful exposure (IRR > 1), the risk ratio is smaller than the IRR, increasingly so as $R_0(t_0, t_1)$ increases, and PoC estimated from the IRR is larger than PoC estimated from the risk ratio. The period enters Equation 4 through $R_0(t_0, t_1)$, which depends on both the length of the period and the background incidence rates (e.g., at the ages it covers); a background risk therefore needs to be specified together with the time origin and end of follow-up to which it applies. When competing risks are not negligible, Equation 4 refers to risks in the absence of competing events, which no longer correspond to the observable cumulative incidence when exposure also increases competing causes of death, as asbestos does. Compensation thresholds in this study were derived from the IRR without applying Equation 4.

**Effect modification: sensitivity analyses**

Between-study heterogeneity in the asbestos exposure-response may reflect differences in study design and exposure assessment, or genuine modification of the exposure-response by characteristics that differ across the source populations, with different implications for a compensation threshold derived from a single pooled estimate. Candidate modifiers were examined as a sensitivity analysis. Each modifier was fitted in a separate model on the same one-stage specification as the main analysis: a mixed-effects logistic regression with random study intercepts, a random cumulative asbestos slope per study, and fixed effects for age group, sex, cigarette pack-years and time since quitting smoking. Modifiers were tested one at a time rather than jointly as centre-level characteristics could be correlated with one another and with study, thus impeding separating their contributions to heterogeneity.

- **Study-level modifiers** were fitted as across-study interactions with the cumulative asbestos slope. Four variables were considered: source of controls (population vs hospital-based); SYN-JEM measurement coverage, defined per participant as the share of their asbestos exposure years falling inside the SYN-JEM measurement window (1971–2009) and averaged over the ever-exposed in each centre; exposure era, defined as the centre median of the first year of asbestos exposure among the ever-exposed; and recruitment era, defined as the midpoint of the recruitment period, reported as a secondary era variable because it indexes follow-up-period background mortality, competing risks and case ascertainment rather than the exposure-response. The two era variables are correlated and neither is identified independently of the other across this number of centres.
- **Individual-level modifiers** included sex, smoking intensity (cigarette pack-years), smoking status, attained age, time since last asbestos exposure and interview year. These variables were decomposed into a within-study and an across-study component before being fitted, following the recommendation for IPD meta-analysis to avoid ecological bias.[17] The covariate was centred about its study mean and the study mean was carried as its own interaction with cumulative asbestos. The within-study coefficient thus estimates individual-level modification free of aggregation bias while the across-study coefficient carries the ecological contrast that a single `asbestos × modifier` interaction would convey. Time since last exposure is undefined for the never-exposed and was set to zero for them, corresponding to cumulative exposure of zero; its contribution to the interaction term is therefore zero whatever value it takes and its study mean was computed over the ever-exposed only, so that the ecological component reflects each centre's exposure timing rather than its exposure prevalence. Interview year was fitted as the deviation from each centre's own mean interview year, with a main effect for interview year included so that the interaction does not absorb the systematic difference in timing between cases and controls; its within-centre component tests the constant-hazard-ratio condition described above.

For each modifier we report a joint Wald test on the modifier × asbestos slope terms, restricted to the within-study terms for individual-level modifiers, and a likelihood ratio test against the corresponding no-interaction model. The likelihood ratio test is omitted where the modifier introduces missing values, because the two models are then not fitted to the same observations. We also report the variance of the random asbestos slope before and after the modifier is included, as the one-stage counterpart of the reduction in between-study heterogeneity for a one-stage meta-analysis (**Supplementary Table 7**). Exposure-response relations, minimum exposures for a PoC of 50% and expected numbers of compensated cases were then estimated per stratum exactly as in the main analysis, for study-level (**Supplementary Table 8**) and individual-level (**Supplementary Table 9**) modifiers.

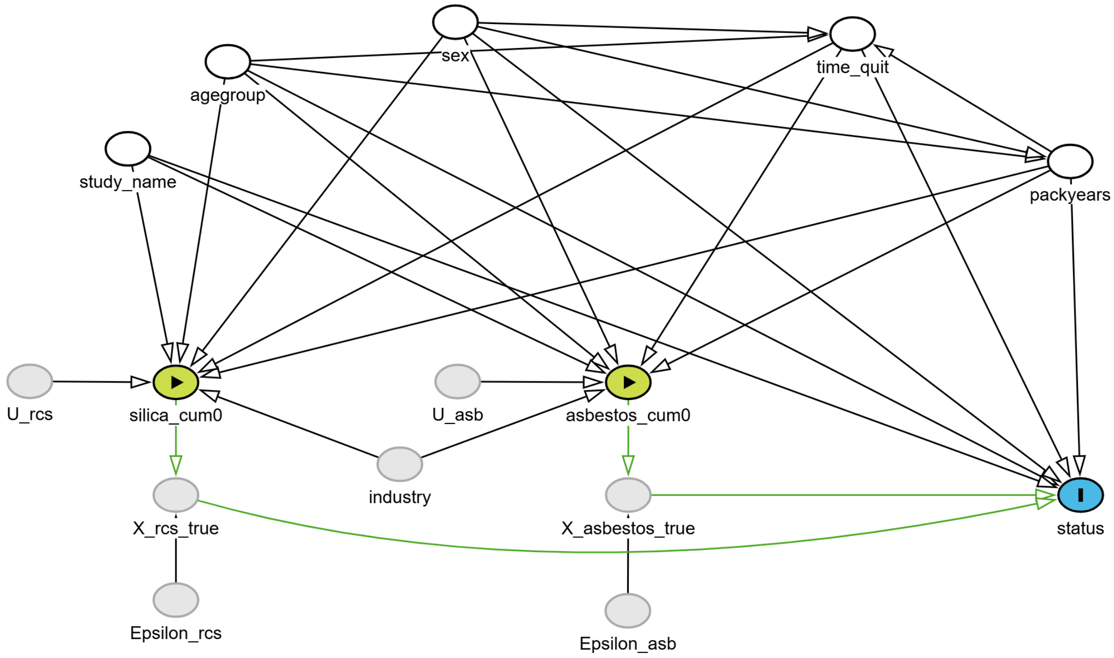

**Supplementary Figure 1.** Directed acyclic graph (DAG) depicting the causal relationship between two occupational lung carcinogens and lung cancer. Each circle (node) represents a variable, while arrows represent the hypothesised relationships between variables. Unobserved variables are coloured in grey, whereas observed variables are green (exposures), blue (outcome) and white (adjusted). The minimal set of adjustment for this model consists of age (agegroup), sex, smoking status and time since quitting (time_quit), lifetime smoking intensity (packyears), and study (study_name). The true value of exposure (X_asb_true and X_rcs_true) are unobserved and thus estimated with a job exposure matrix (asbestos_cum0 and silica_cum0), resulting in loss of individual variation (Epsilon), and exposure error terms (U) that are independent of outcome status (Berkson-type error). This DAG and its associated model code can be accessed through OpenCausal.org https://opencausal.org/graph/bwoy9fng and dagitty https://dagitty.net/mjsrVdmjg.

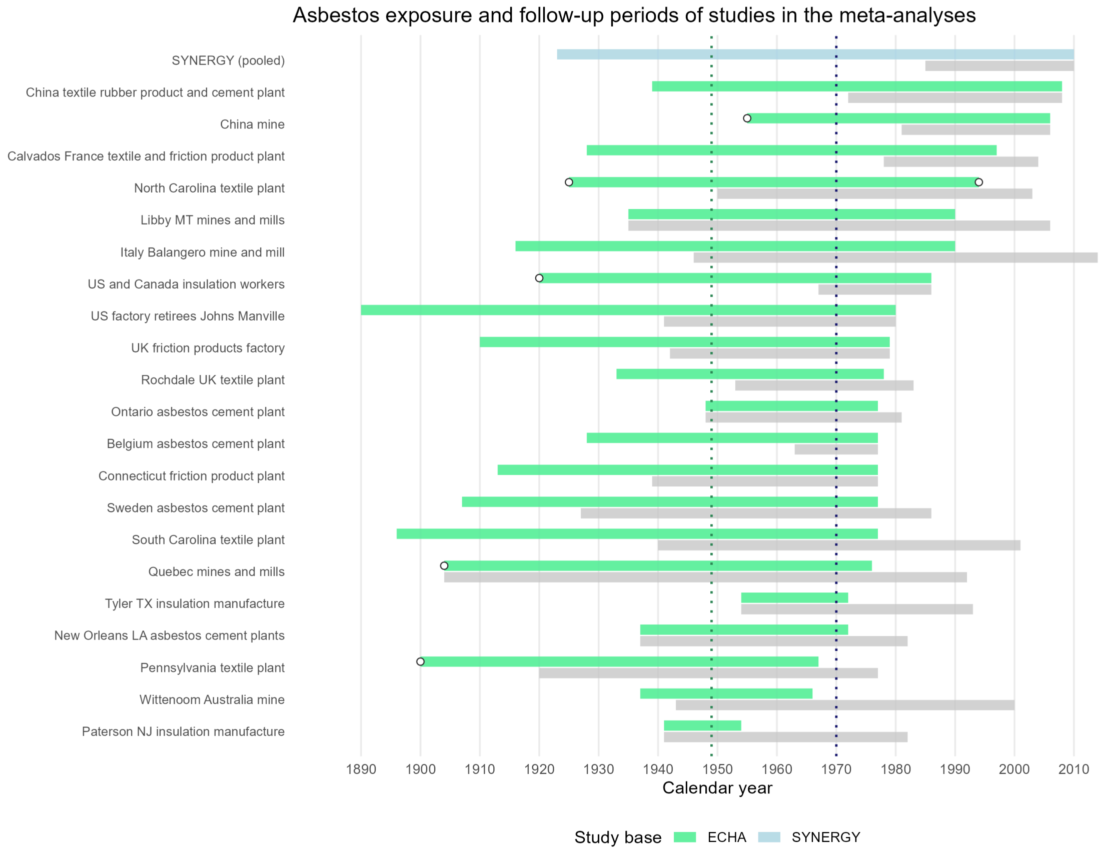

**Supplementary Figure 2.** Calendar periods of asbestos exposure and follow-up of the studies in the European Chemicals Agency (ECHA) study base, with the SYNERGY pooled case-control study shown as a single study. For each study, the upper bar represents the asbestos exposure period (green: occupational studies; blue: SYNERGY) and the lower bar (grey) the follow-up period (for SYNERGY, the period of case and control recruitment). Open circles mark exposure start or end years reported as approximate in the source publications. Dotted vertical lines mark the calendar year by which half of the exposure-years had elapsed in the occupational studies (green; exposure periods expanded year by year and weighted by study size) and in SYNERGY (dark blue; individual exposure-years). Exposure and follow-up periods of the occupational studies were obtained from Lenters, et al.[18] and from the original publications for the studies added[19–21] or updated by ECHA.[22,23] The exposure period of SYNERGY was obtained from individual participant data.

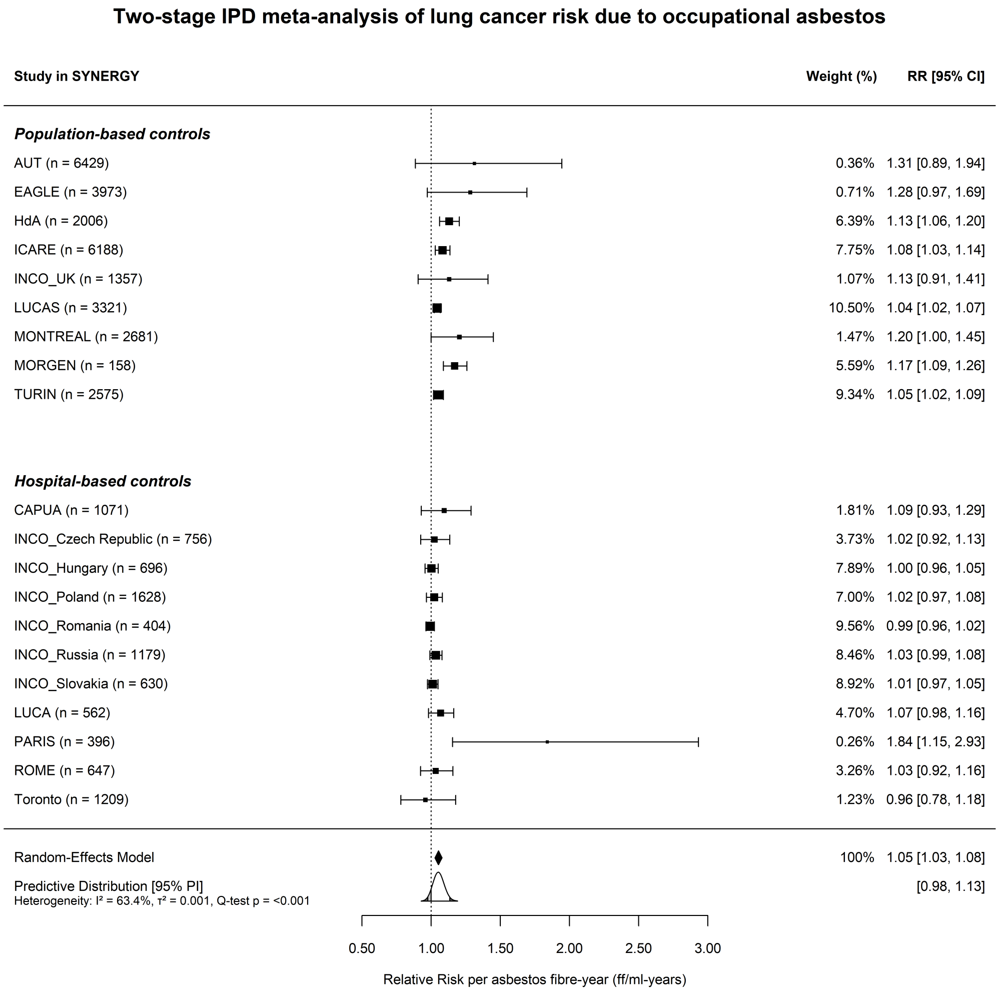

**Supplementary Figure 3.** Forest plot of study-specific and pooled effect estimates of the relative risk of lung cancer per asbestos fibre-year in SYNERGY. The INCO Poland and Toronto studies included both population and hospital-based controls, and are considered as hospital-based controlled studies for this analysis for consistency with prior SYNERGY meta-analyses.

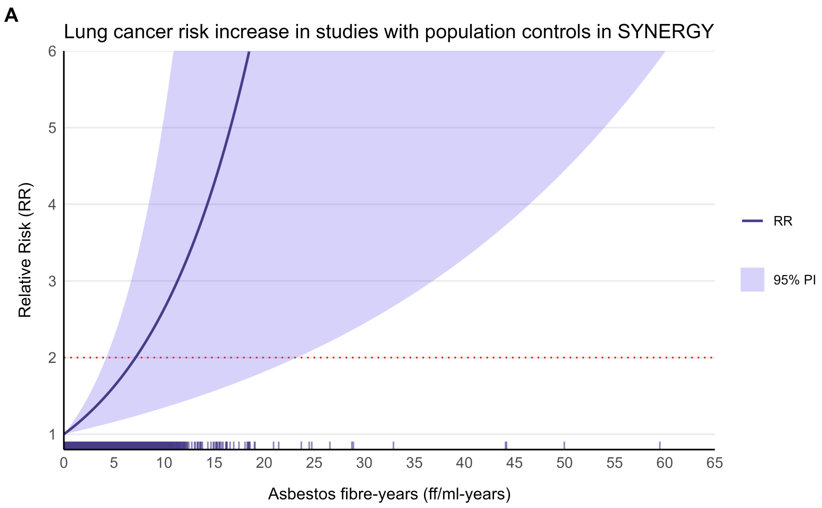

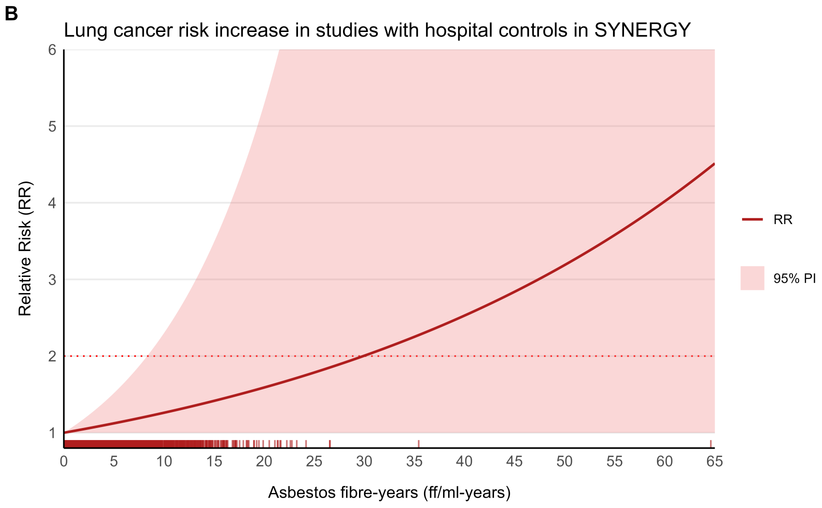

**Supplementary Figure 4.** Exposure response relations of lung cancer risk increase at low cumulative asbestos exposure in the SYNERGY study, stratified according to the source of controls: population controls **A)** and hospital controls **B)**. Models were adjusted for age, sex, smoking (pack-years), and time since quitting smoking. Solid lines represent the average meta-estimates; shaded areas, the 95% prediction intervals (95% PI), and the dotted red line, the doubling in increase of risk threshold (RR = 2). Vertical tick marks represent observed individual participant exposure values in each subset.

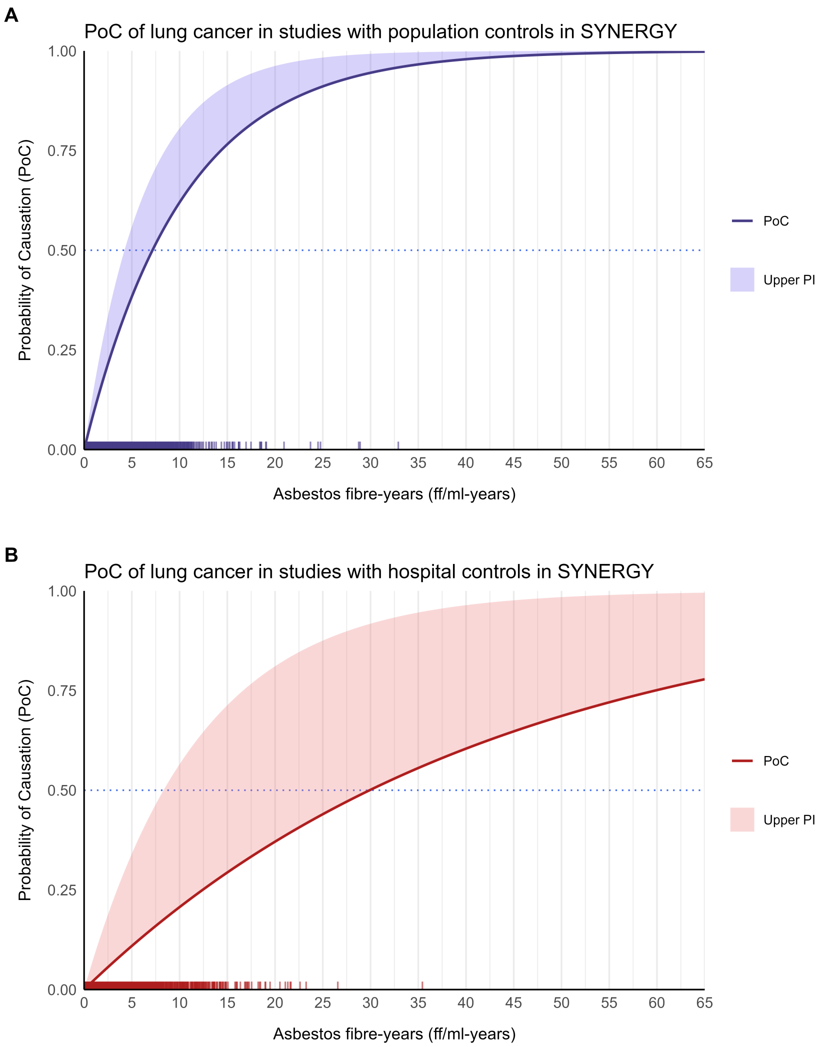

**Supplementary Figure 5.** Probability of causation of lung cancer due to cumulative asbestos exposure in the SYNERGY study, stratified according to the source of controls: population controls **A)** and hospital controls **B)**. Models were adjusted for age, sex, smoking (pack-years), and time since quitting smoking. Solid lines represent the average meta-estimates; shaded areas, the upper 95% prediction interval (Upper PI), and the dotted blue line, the PoC = 0.5 threshold. Vertical tick marks represent observed individual participant exposure values of lung cancer cases in each subset.

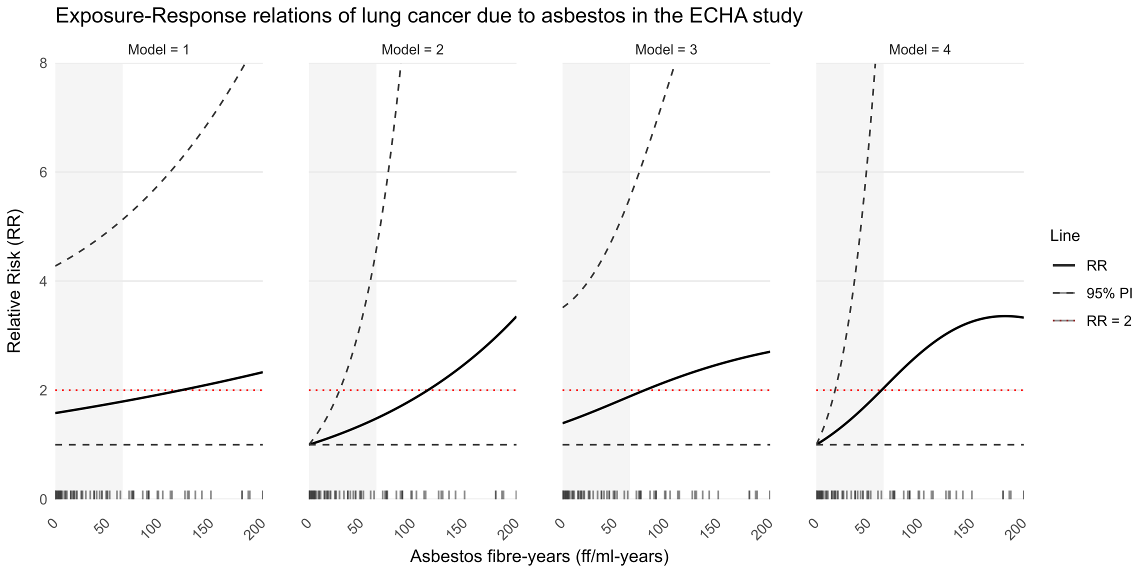

**Supplementary Figure 6.** Exposure-response relations under different model assumptions: (1) linear model, assumes difference in background rate of outcome; (2) linear model, assumes no difference in background rate of outcome; (3) spline model, assumes difference in background rate of outcome; (4) spline model, assumes no difference in background rate of outcome. The models were fitted using logRR estimates from the studies reviewed by Lenters, et al.[18] and van der Bij, et al.[6], updated in 2021 for the European Chemicals Agency report on occupational exposure limits for asbestos.[24] The grey-shaded area represents the range of exposure values observed in SYNERGY. The horizontal red dashed line marks the relative risk threshold at which RR = 2. Vertical tick marks (bottom) represent average exposure values for the reported pooled estimates per exposure category in ECHA. Abbreviations: Relative risk (RR), 95% prediction interval (PI).

**Supplementary Table 1.** Exposure assessment method in the 22 studies included in the European Chemicals Agency (ECHA) study base, including the SYNERGY pooled case-control study.

| Study | Cumulative asbestos estimation method | Measurement Method (Period) | Coverage, % [a] | Conversion factor [b] | JEM assignment level | Job History completeness [c] |
|---|---|---|---|---|---|---|
| Quebec, Canada, mines and mills[25] | JEM | Impinger (1948-1966), PCM (1969-1976) | 25 | Internal | Job, 1-year | Insufficient |
| Italy, Balangero, mine and mill[22] | JEM | PCM (1969-1990) | 24 | N/A | Job, time resolution not reported | Sufficient |
| Connecticut, friction product plant[26] | JEM | Impinger (1930,35,36,39), PCM (1970-1977) | 30 | External | Department, 10-year | Insufficient |
| South Carolina, textile plant[27] | JEM | Impinger (1930-1971), PCM (1965-1977) | 58 | Internal | Operation, department, 1-year | Sufficient |
| North Carolina, textile plant[28] | JEM | Impinger (1935-1971), PCM (1964-1986) | 74 | Internal | Job, department, plant, >10-year periods | Insufficient |
| Wittenoom, Australia, mine[29] | JEM | Coniometer (1948-1958), PCM (1966) | 5 | N/A | Job/work-area, time resolution not reported | Sufficient |
| Patterson, NJ, insulation manufacture[30] | JEM (external reference) | None | 0 | N/A | Job, single external exposure estimate (no time resolution) | Sufficient |
| Tyler, TX, insulation manufacture[31] | Years of exposure x 45 fibres/ml (plant average) | PCM (1967,70,71) | 25 | N/A | N/A | Sufficient |
| Libby, MT, mines and mills[23] | JEM | Impinger (1956-1969), PCM (1967-1982) | 47 | Internal | Job, 10-year (pre-1970, not reported thereafter) | Sufficient |
| UK, friction products factory[32] | JEM | PCM (1967-1979) | 19 | N/A | Cost-centre, time resolution not reported | Sufficient |
| Ontario, asbestos cement plant[33] | JEM | Impinger (1949,54-57), PCM (1969-1977) | 80 | External | Job, time resolution not reported | Insufficient |

| Study | Cumulative asbestos estimation method | Measurement Method (Period) | Coverage, % [a] | Conversion factor [b] | JEM assignment level | Job History completeness [c] |
|---|---|---|---|---|---|---|
| New Orleans, LA, asbestos cement plants[34] | JEM | Impinger (1952-1969), PCM (1969-1972) | 61 | Internal | Job, time resolution not reported | Insufficient |
| Sweden, asbestos cement plant[35] | JEM | Impinger/gravimetric (1956-1969), PCM (1969-1977) | 30 | Internal | Job, 5-year | Insufficient |
| Belgium, asbestos cement plant[36] | JEM | PCM (1970-1976) | 12 | N/A | Work area, time resolution not reported | Sufficient |
| US factory retirees (Johns Manville)[37] | JEM | Impinger (Mid-1950s-1980) | 30 | External | Job, time resolution not reported | Sufficient |
| US and Canada, insulation workers[38] | Years from exposure onset x 15 fibres/ml | None | 0 | External | N/A | Sufficient |
| Pennsylvania, textile plant[39] | JEM | Impinger (1930-39,56-67), PCM (1967) | 55 | External | Department, 10-year | Sufficient |
| Rochdale, UK, textile plant[40] | JEM | Impinger (1951-1964), PCM (1965-1978) | 60 | Internal | Job exposure category (low, medium, high, very high), 5-year | Sufficient |
| Calvados, France, textile and friction product plant[19] | JEM | Gravimetric (1959-1973), PCM (1973-1997) | 60 | Internal | Workshop, plant, 1-year | Sufficient |
| China, textile, rubber product and cement plant[20] | JEM | Gravimetric (1955–1990), PCM (1999,2002,2006), EM (2006) | 77 | Internal | Job, work area, 5-year | Sufficient |
| China, mine[21] | JEM | Gravimetric (1984-1995,2006), PCM (1991,2006) | 40 | Internal | Job, workshop, study duration-averaged | Sufficient |

| Study | Cumulative asbestos estimation method | Measurement Method (Period) | Coverage, % [a] | Conversion factor [b] | JEM assignment level | Job History completeness [c] |
|---|---|---|---|---|---|---|
| Europe and Canada pooled case-control study[41] | JEM (SYN-JEM) | >95% PCM and <5% EM (1971-2009) | 49 | N/A | Job, region, 1-year | Sufficient |

Abbreviations: electron microscopy (EM), job exposure matrix (JEM), phase contrast microscopy (PCM).

[a] Coverage as evaluated by Lenters, et al.[18]: "Percentage of the accumulated work history years temporally covered by exposure measurement data provides an indication of the extent temporal back-extrapolation or reconstruction approaches were used in the exposure estimation process"

[b] Numerical factors used to convert particles measured with older methods (e.g. impinger) to fibres measured with microscopy (e.g, PCM and EM). Internal factors were obtained in validation subsets of the same cohort, whereas external conversion factors used those obtained in different study populations.

[c] Job history completeness as evaluated by Lenters, et al.[18]: "We considered whether the job history information was sufficiently complete and detailed to capture changes in job titles or tasks over time and between companies, and sufficiently refined or appropriately used in a way that captured differences in exposure between jobs."

**Supplementary Table 2.** Study base and matching design of the SYNERGY pooled case-control study centres.

| Study | Study base | Matching type | Matching variables |
|---|---|---|---|
| AUT[42] | Dynamic population | Individual + frequency[a] | Sex, age, region |
| CAPUA[43] | Dynamic population | Individual[b] | Ethnicity, sex, age (±5 y) |
| EAGLE[44] | Dynamic population | Frequency | Age (5-y groups), sex, area (5 areas) |
| HdA[45] | Dynamic population | Individual[a] | Region, sex, age (±5 y) |
| ICARE[46] | Dynamic population | Frequency | Age, sex, département[c] |
| INCO Czech Republic[47,48] | Dynamic population | Frequency | Age (±3 y), sex, study area |
| INCO Hungary[47,48] | Dynamic population | Frequency | Age (±3 y), sex, study area |
| INCO Poland[47,48] | Dynamic population | Frequency | Age (±3 y), sex, study area |
| INCO Romania[47,48] | Dynamic population | Frequency | Age (±3 y), sex, study area |
| INCO Russia[47,48] | Dynamic population | Frequency | Age (±3 y), sex, study area |
| INCO Slovakia[47,48] | Dynamic population | Frequency | Age (±3 y), sex, study area |
| INCO UK[48] | Dynamic population | Frequency | Age (±3 y), sex, study area |
| LUCA[8] | Dynamic population | Individual[d] | Age (±2.5 y), hospital |
| LUCAS[9] | Dynamic population | Frequency | Age (5-y groups), year of inclusion |
| MONTREAL[49] | Dynamic population | Frequency | Sex, age |
| MORGEN[11] | Cohort (nested) | NA[e] | NA[e] |
| PARIS[50] | Dynamic population | Frequency | Age, sex, hospital |
| ROME[51] | Dynamic population | Frequency | Sex, age (5-y groups) |
| Toronto[10] | Dynamic population | Frequency | Sex, ethnicity |
| TURIN[52] | Dynamic population | Frequency | Sex, age (5-y groups) |

Abbreviation: y, years.

[a] AUT pools two sub-studies: BIPS (individually matched, 1:1) and the GSF radon study (frequency matched). The pooled analysis conditioned on age (6 categories) and region (17 categories) strata rather than on the original matched sets. The BIPS component is the study reported separately as HdA.

[b] The Methods describe individual matching and the Results refer to frequency matching; the analysis was unconditional and excluded 194 cases without a matched control.

[c] Controls were additionally stratified on socioeconomic status (conditional on sex), based on last job held.

[d] Controls were recruited concurrently, one sought for each incident case, but calendar time was not a stated matching variable.

[e] The cohort profile does not document a nested case-control sampling scheme, matching, or control selection specifically for lung cancer.

**Supplementary Table 3.** Cumulative asbestos exposure among cases and controls per study in SYNERGY, ordered by source of controls (hospital and population-based control studies).

| Source Controls | Study | Exposed (%) | | Median exposure (ff/ml-years) (among exposed) | | IQR (ff/ml-years) (among exposed) | | Range (ff/ml-years) (among exposed) | |
|---|---|---|---|---|---|---|---|---|---|
| | | *Cases* | *Controls* | *Cases* | *Controls* | *Cases* | *Controls* | *Cases* | *Controls* |
| H | CAPUA[43] | 42.0 | 39.3 | 1.493 | 1.595 | 0.691 - 2.685 | 0.871 - 2.587 | 0.013 - 16.913 | 0.018 - 10.055 |
| H | INCO Czech Republic[47,48] | 44.4 | 34.3 | 3.800 | 4.328 | 1.713 - 6.611 | 1.746 - 6.852 | 0.101 - 35.432 | 0.112 - 15.412 |
| H | INCO Hungary[47,48] | 50.4 | 45.2 | 3.866 | 4.182 | 1.43 - 7.867 | 1.946 - 7.19 | 0.042 - 21.661 | 0.107 - 64.6 |
| H | INCO Poland[47,48] | 35.1 | 29.3 | 2.850 | 2.364 | 1.195 - 5.947 | 1.06 - 5.535 | 0.018 - 21.593 | 0.025 - 24.183 |
| H | INCO Romania[47,48] | 33.0 | 24.4 | 3.287 | 3.076 | 1.809 - 6.82 | 1.432 - 5.554 | 0.141 - 15.954 | 0.053 - 15.853 |
| H | INCO Russia[47,48] | 55.4 | 54.1 | 3.400 | 3.839 | 1.926 - 6.962 | 1.766 - 7.307 | 0.048 - 26.571 | 0.03 - 26.55 |
| H | INCO Slovakia[47,48] | 43.5 | 34.7 | 3.631 | 3.570 | 1.442 - 7.323 | 1.43 - 7.104 | 0.145 - 23.255 | 0.162 - 17.194 |
| H | LUCA[8] | 43.6 | 39.0 | 1.060 | 1.351 | 0.484 - 2.265 | 0.559 - 2.286 | 0.053 - 6.933 | 0.049 - 6.535 |
| H | PARIS[53] | 41.4 | 45.4 | 2.144 | 1.664 | 0.99 - 3.167 | 0.823 - 3.204 | 0.072 - 7.035 | 0.123 - 7.547 |
| H | ROME[51] | 46.3 | 38.0 | 2.744 | 2.502 | 1.12 - 3.732 | 0.908 - 3.736 | 0.131 - 7.673 | 0.022 - 9.367 |
| H | Toronto[10] | 21.4 | 10.5 | 0.466 | 0.196 | 0.23 - 1.008 | 0.071 - 0.555 | 0.033 - 2.378 | 0.006 - 2.27 |
| P | AUT[42] | 53.2 | 41.2 | 2.576 | 1.979 | 1.028 - 5.049 | 0.975 - 4.512 | 0.007 - 24.758 | 0.035 - 59.509 |
| P | EAGLE[44] | 35.0 | 26.5 | 1.451 | 1.261 | 0.602 - 2.67 | 0.54 - 2.561 | 0.013 - 8.361 | 0.017 - 10.853 |
| P | HdA[45] | 56.5 | 45.6 | 2.618 | 2.389 | 1.034 - 5.09 | 1.058 - 4.909 | 0.104 - 32.909 | 0.045 - 21.454 |
| P | ICARE[46] | 41.2 | 32.7 | 0.869 | 0.696 | 0.379 - 1.634 | 0.294 - 1.391 | 0.002 - 7.599 | 0.002 - 15.236 |
| P | INCO UK[48] | 61.2 | 52.2 | 0.345 | 0.328 | 0.153 - 0.65 | 0.16 - 0.643 | 0.011 - 5.889 | 0.004 - 4.43 |
| P | LUCAS[9] | 28.9 | 26.6 | 0.628 | 0.549 | 0.324 - 0.858 | 0.242 - 0.859 | 0.021 - 2.117 | 0.003 - 1.803 |
| P | MONTREAL[49] | 36.8 | 30.5 | 0.575 | 0.531 | 0.247 - 1.11 | 0.222 - 1.067 | 0.013 - 4.94 | 0.003 - 3.655 |
| P | MORGEN[11] | 27.9 | 10.4 | 1.913 | 2.509 | 1.265 - 2.674 | 1.546 - 4.141 | 0.471 - 4.617 | 0.404 - 6.942 |
| P | TURIN[52] | 52.6 | 43.4 | 2.075 | 1.742 | 0.944 - 3.677 | 0.889 - 3.492 | 0.032 - 19.084 | 0.055 - 11.557 |

Abbreviations: hospital-based controls (H), interquartile range (IQR), population-based controls (P), percentage (%).

**Supplementary Table 4.** Smoking among cases and controls per study in SYNERGY, ordered by source of controls (hospital and population-based control studies).

| Source Controls | Study | Ever-smoker (%) | | Median pack-years (among exposed) | | IQR (among exposed) | | Range (among exposed) | |
|---|---|---|---|---|---|---|---|---|---|
| | | *Cases* | *Controls* | *Cases* | *Controls* | *Cases* | *Controls* | *Cases* | *Controls* |
| H | CAPUA[43] | 95.0 | 68.4 | 54.0 | 31.5 | 39 - 76.2 | 13.2 - 52.8 | 0.7 - 240 | 0 - 170 |
| H | INCO Czech Republic[47,48] | 92.1 | 57.3 | 38.5 | 20.0 | 26.5 - 49.8 | 9.7 - 36.8 | 0.3 - 152 | 0 - 141.2 |
| H | INCO Hungary[47,48] | 91.0 | 65.2 | 35.0 | 29.5 | 24.5 - 46.6 | 15.2 - 41.5 | 2 - 115.5 | 0.3 - 144.4 |
| H | INCO Poland[47,48] | 94.7 | 70.8 | 37.2 | 27.2 | 28 - 48.9 | 15.5 - 38 | 2 - 127.6 | 0 - 120.7 |
| H | INCO Romania[47,48] | 88.8 | 54.7 | 34.5 | 20.9 | 22.1 - 46.7 | 10.6 - 32.5 | 1 - 138.2 | 0.7 - 129.4 |
| H | INCO Russia[47,48] | 88.0 | 73.6 | 31.5 | 23.4 | 22.3 - 40.5 | 13.2 - 34.2 | 0.3 - 99 | 0.3 - 90.9 |
| H | INCO Slovakia[47,48] | 93.0 | 60.4 | 36.0 | 25.6 | 25.8 - 47 | 14.1 - 39.1 | 0.8 - 131.8 | 0.3 - 141 |
| H | LUCA[8] | 99.6 | 81.9 | 37.0 | 26.5 | 25.8 - 49 | 11.4 - 40.2 | 1.5 - 111 | 0.1 - 140 |
| H | PARIS[53] | 99.4 | 97.8 | 19.0 | 43.6 | 11.6 - 32.2 | 30.6 - 63.8 | 1.6 - 58 | 3 - 232.7 |
| H | ROME[51] | 93.3 | 74.5 | 51.4 | 35.4 | 35 - 76.4 | 17.1 - 50.9 | 0.6 - 180 | 0.2 - 191 |
| H | Toronto[10] | 59.7 | 43.5 | 42.0 | 17.5 | 23 - 60 | 6.5 - 36.2 | 0.3 - 235 | 0.1 - 220 |
| P | AUT[42] | 92.7 | 67.0 | 32.5 | 17.6 | 21.5 - 44.5 | 7.2 - 30.5 | 0 - 145.8 | 0 - 188.9 |
| P | EAGLE[44] | 93.2 | 67.9 | 44.8 | 23.5 | 30.8 - 60 | 9.6 - 40 | 0 - 315 | 0 - 211.5 |
| P | HdA[45] | 90.2 | 70.0 | 31.5 | 21.0 | 20.6 - 43.5 | 10 - 33.5 | 0.3 - 167.4 | 0 - 136 |
| P | ICARE[46] | 91.6 | 61.0 | 36.2 | 16.1 | 24.2 - 50.2 | 6.6 - 28 | 0.1 - 161.8 | 0 - 140 |
| P | INCO UK[48] | 93.9 | 71.7 | 38.4 | 24.0 | 25.4 - 52 | 11.8 - 38 | 0.3 - 252 | 0 - 117 |
| P | LUCAS[9] | 88.0 | 62.5 | 38.0 | 24.0 | 23.4 - 52 | 9.6 - 37.4 | 0.2 - 105 | 0.2 - 94.5 |
| P | MONTREAL[49] | 95.6 | 68.5 | 58.8 | 38.8 | 45 - 85 | 18.8 - 60 | 0.8 - 285 | 0 - 318.8 |
| P | MORGEN[11] | 95.3 | 47.8 | 30.0 | 22.0 | 22 - 40 | 14 - 30.2 | 3.9 - 90.7 | 3.6 - 45.8 |
| P | TURIN[52] | 94.7 | 71.5 | 36.8 | 25.0 | 24.8 - 48 | 13.5 - 39.2 | 0.2 - 182 | 0.1 - 139.8 |

Abbreviations: hospital-based controls (H), interquartile range (IQR), population-based controls (P), percentage (%).

**Supplementary Table 5.** Comparison of exposure-response relations of lung cancer risk and asbestos in the meta-regression of published occupational asbestos studies including and excluding the SYNERGY study.

| Model | Risk increase per fibre-year[a] | | Min exposure for 50% PoC (fibre-years)[b] | | Cases per 10,000 above 50% PoC in SYNERGY | | Heterogeneity ($I^2$) |
|---|---|---|---|---|---|---|---|
| | Estimate | 95% Prediction Interval | Point estimate | Presumably plausible | Point estimate | Presumably plausible | |
| **Meta-regression of 21 occupational asbestos studies excluding SYNERGY** | | | | | | | |
| Spline (21 studies, 90,823 participants) | 0.9% | [0%, 2.5%] | 84.05 | 26.56 | 0 | 3 | 92.9 |
| **Meta-regression of 22 occupational asbestos studies including SYNERGY** | | | | | | | |
| Spline (22 studies, 130,341 participants) | 1.1% | [0%, 3.6%] | 62.93 | 18.19 | 0 | 17 | 92.7 |

Abbreviations: probability of causation (PoC).

[a] Negative values truncated to 0 due to monotonicity assumption. For spline model: instantaneous slope at 1.5 fibre-years (median exposure in cases).

[b] A probability of causation (PoC) of 50% corresponds to a doubling in lung cancer risk (i.e., RR = 2) for the point estimate, and the upper 95% prediction interval for the presumably plausible threshold.

**Supplementary Table 6.** Comparison of exposure-response relations of lung cancer risk and cumulative asbestos exposure in the SYNERGY study under different individual participant data meta-analyses, overall and stratified by source of controls.

| Method[c] | Risk increase per fibre-year[a] | | Min exposure for 50% PoC (fibre-years)[b] | | Cases per 10,000 above 50% PoC in SYNERGY | |
|---|---|---|---|---|---|---|
| | Estimate | 95% Prediction Interval | Point estimate | Presumably plausible | Point estimate | Presumably plausible |
| **Main meta-analysis** | | | | | | |
| Two-stage univariate | 5.2% | [0%, 13.5%] | 13.61 | 5.49 | 47 | 581 |
| One-stage stratified | 5.3% | [0%, 13.6%] | 13.34 | 5.42 | 52 | 590 |
| One-stage selective random-effects | 6.8% | [0%, 17.5%] | 10.52 | 4.30 | 117 | 846 |
| **Population-based control studies** | | | | | | |
| Two-stage univariate | 9.6% | [0.3%, 19.7%] | 7.57 | 3.86 | 307 | 978 |
| One-stage stratified | 8.3% | [2.9%, 14.0%] | 8.67 | 5.29 | 204 | 615 |
| One-stage selective random-effects | 10.2% | [3.7%, 17.0%] | 7.16 | 4.41 | 344 | 820 |
| **Hospital-based control studies** | | | | | | |
| Two-stage univariate | 1.3% | [0%, 3.1%] | 53.29 | 23.04 | 0 | 5 |
| One-stage stratified | 1.8% | [0%, 7.0%] | 38.18 | 10.26 | 0 | 128 |
| One-stage selective random-effects | 2.3% | [0%, 8.6%] | 29.92 | 8.40 | 1 | 223 |

Abbreviations: probability of causation (PoC).

[a] Negative values truncated to 0 due to monotonicity assumption.

[b] A probability of causation (PoC) of 50% corresponds to a doubling in lung cancer risk (i.e., RR = 2) for the point estimate, and the upper 95% prediction interval for the presumably plausible threshold.

[c] Two-stage univariate: traditional random-effects meta-analysis of exposure coefficients. One-stage stratified: mixed-effects logistic regression model with stratified intercepts per study, random cumulative asbestos slopes within study, and fixed effects for confounders (age group, sex, time since quitting smoking, and cigarette pack-years). One-stage selective random-effects: mixed-effects logistic regression model with random study intercepts, random cumulative asbestos slopes within study, and fixed effects for confounders (age group, sex, time since quitting smoking, and cigarette pack-years).

**Supplementary Table 7.** Effect modification of the cumulative asbestos exposure-response for lung cancer and one-stage heterogeneity, by modifier, in the SYNERGY study.

| **Modifier**[a] | **Effect-modification test**[b] | | | | | **One-stage random-slope variance**[c] | |
|---|---|---|---|---|---|---|---|
| | **Wald df** | **Wald $\chi^2$** | **Wald *p*** | **LRT df** | **LRT *p*** | **$\tau^2$ base** | **$\tau^2$ modelled** |
| Source of controls | 1 | 16.93 | <0.001 | 2 | 0.002 | $2.18 \times 10^{-3}$ | $7.69 \times 10^{-4}$ |
| JEM measurement coverage | 1 | 0.34 | 0.559 | 2 | 0.638 | $2.18 \times 10^{-3}$ | $2.11 \times 10^{-3}$ |
| Exposure era | 1 | 0.00 | 0.947 | 2 | 0.593 | $2.18 \times 10^{-3}$ | $2.28 \times 10^{-3}$ |
| Recruitment era (secondary) | 1 | 0.36 | 0.550 | 2 | 0.422 | $2.18 \times 10^{-3}$ | $2.32 \times 10^{-3}$ |
| Sex | 1 | 1.63 | 0.202 | 2 | 0.159 | $2.18 \times 10^{-3}$ | $2.00 \times 10^{-3}$ |
| Smoking intensity | 1 | 0.21 | 0.646 | 2 | 0.123 | $2.18 \times 10^{-3}$ | $2.69 \times 10^{-3}$ |
| Smoking status | 2 | 6.40 | 0.041 | 4 | <0.001 | $2.18 \times 10^{-3}$ | $5.10 \times 10^{-4}$ |
| Attained age | 1 | 3.13 | 0.077 | 2 | 0.146 | $2.18 \times 10^{-3}$ | $1.80 \times 10^{-3}$ |
| Time since last exposure | 1 | 0.23 | 0.630 | —[d] | —[d] | $2.18 \times 10^{-3}$ | $1.60 \times 10^{-3}$ |
| Interview year (within centre) | 1 | 2.15 | 0.143 | —[d] | —[d] | $2.18 \times 10^{-3}$ | $2.14 \times 10^{-3}$ |

Abbreviations: degrees of freedom (df), job exposure matrix (JEM), likelihood ratio test (LRT).

[a] All models use random intercepts per study. Every modifier is fitted marginally, so the baseline for each row is the no-modifier model on the same random structure. Study-level modifiers enter as across-study interactions; individual-level modifiers (sex, smoking intensity, smoking status, attained age, time since last exposure, interview year) are fitted with their within- and across-study components separated, and only the within-study component is tested here.

[b] Joint Wald test on the modifier × asbestos slope terms, and likelihood ratio test against the corresponding no-interaction model.

[c] $\tau^2$ is the variance of the random asbestos slope in the one-stage model, before (base) and after (modelled) the modifier is included.

[d] The likelihood ratio test is not reported when the modifier introduces missing values, because the two models are then not fitted to the same observations.

**Supplementary Table 8.** Comparison of exposure-response relations of lung cancer risk and cumulative asbestos exposure in the SYNERGY study, by study-level modifier, under the one-stage model with random intercepts.

| Model | Risk increase per fibre-year[a] | | Min exposure for 50% PoC (fibre-years)[b] | | Cases per 10,000 above 50% PoC in SYNERGY | |
|---|---|---|---|---|---|---|
| | Estimate | 95% Prediction Interval | Point estimate | Presumably plausible | Point estimate | Presumably plausible |
| **No modifier (reference)** | | | | | | |
| One-stage, random intercepts | 6.8% | [0%, 17.5%] | 10.52 | 4.30 | 117 | 846 |
| **JEM measurement coverage** | | | | | | |
| 30% | 7.6% | [0%, 18.7%] | 9.43 | 4.05 | 160 | 916 |
| 45% | 6.7% | [0%, 17.2%] | 10.71 | 4.37 | 111 | 830 |
| 55% | 6.0% | [0%, 16.8%] | 11.83 | 4.47 | 75 | 805 |
| **Exposure era** | | | | | | |
| 1954 | 6.9% | [0%, 18.0%] | 10.39 | 4.18 | 122 | 875 |
| 1958 | 6.8% | [0%, 17.8%] | 10.49 | 4.24 | 118 | 864 |
| 1963 | 6.7% | [0%, 18.1%] | 10.63 | 4.17 | 114 | 878 |
| **Recruitment era (secondary)** | | | | | | |
| 1992 | 5.9% | [0%, 17.3%] | 12.10 | 4.33 | 69 | 841 |
| 1999 | 7.2% | [0%, 18.3%] | 9.99 | 4.12 | 139 | 893 |
| 2000 | 7.3% | [0%, 18.6%] | 9.78 | 4.07 | 146 | 908 |

Abbreviations: job exposure matrix (JEM), probability of causation (PoC).
[a] Negative values truncated to 0 due to monotonicity assumption.
[b] A probability of causation (PoC) of 50% corresponds to a doubling in lung cancer risk (i.e., RR = 2) for the point estimate, and the upper 95% prediction interval for the presumably plausible threshold.

**Supplementary Table 9.** Comparison of exposure-response relations of lung cancer risk and cumulative asbestos exposure in the SYNERGY study, by individual-level modifier, under the one-stage model with random intercepts.

| Model[c] | Risk increase per fibre-year[a] | | Min exposure for 50% PoC (fibre-years)[b] | | Cases per 10,000 above 50% PoC in SYNERGY | |
|---|---|---|---|---|---|---|
| | Estimate | 95% Prediction Interval | Point estimate | Presumably plausible | Point estimate | Presumably plausible |
| **Sex** | | | | | | |
| Female | 10.4% | [0%, 22.7%] | 6.99 | 3.39 | 360 | 1147 |
| Male | 6.6% | [0%, 16.8%] | 10.88 | 4.46 | 102 | 807 |
| **Smoking intensity** | | | | | | |
| 0 pack-years | 8.6% | [0%, 21.1%] | 8.37 | 3.62 | 227 | 1070 |
| 17 pack-years | 8.0% | [0%, 20.1%] | 9.00 | 3.79 | 187 | 1000 |
| 31 pack-years | 7.9% | [0%, 19.9%] | 9.14 | 3.81 | 179 | 994 |
| 46 pack-years | 7.8% | [0%, 19.9%] | 9.23 | 3.83 | 169 | 992 |
| **Smoking status** | | | | | | |
| Never smoker | 6.9% | [0.9%, 13.2%] | 10.45 | 5.58 | 120 | 563 |
| Former smoker | 7.0% | [1.9%, 12.4%] | 10.20 | 5.91 | 131 | 503 |
| Current smoker | 10.1% | [4.7%, 15.8%] | 7.21 | 4.74 | 339 | 732 |
| **Attained age** | | | | | | |
| 55 years | 8.3% | [0%, 18.4%] | 8.66 | 4.11 | 204 | 895 |
| 65 years | 6.9% | [0%, 16.6%] | 10.41 | 4.50 | 122 | 797 |
| 75 years | 5.5% | [0%, 15.2%] | 13.04 | 4.89 | 56 | 695 |
| **Time since last exposure** | | | | | | |
| 5 years | 7.5% | [0%, 16.7%] | 9.63 | 4.48 | 154 | 804 |
| 15 years | 7.7% | [0%, 17.0%] | 9.30 | 4.41 | 166 | 820 |
| 30 years | 8.1% | [0%, 17.7%] | 8.86 | 4.25 | 193 | 862 |

Abbreviations: probability of causation (PoC).
[a] Negative values truncated to 0 due to monotonicity assumption.

[b] A probability of causation (PoC) of 50% corresponds to a doubling in lung cancer risk (i.e., RR = 2) for the point estimate, and the upper 95% prediction interval for the presumably plausible threshold.
[c] Each modifier is fitted in a separate model with its within-study and across-study components separated: the modifier is centred about its study mean, and the study mean is carried as its own interaction with cumulative asbestos exposure. The strata shown are the within-study (individual-level) contrasts, with the across-study component held at the grand mean.

### References

1 Mancilla Galindo J. Causal relationship between two occupational carcinogens and lung cancer. 2025.

https://opencausal.org/graph/bwoy9fng

(accessed 5 Dec 2025).

2 Naimi AI, Cole SR, Hudgens MG, *et al.* Assessing the component associations of the healthy worker survivor bias: occupational asbestos exposure and lung cancer mortality. *Annals of Epidemiology* 2013;**23**:334–41. doi:https://doi.org/10.1016/j.annepidem.2013.03.013

3 Armstrong BG. Effect of Measurement Error on Epidemiological Studies of Environmental and Occupational Exposures. *Occupational and Environmental Medicine* 1998;**55**:651–6. doi:10.1136/oem.55.10.651

4 Olsson A, Bouaoun L, Schüz J, *et al.* Lung Cancer Risks Associated with Occupational Exposure to Pairs of Five Lung Carcinogens: Results from a Pooled Analysis of Case-Control Studies (SYNERGY). *Environmental Health Perspectives* 2024;**132**:17005. doi:10.1289/ehp13380

5 Kamenetsky ME, Keil AP. (Re-)match: adjusting for matching factors in case-control studies may be unnecessary or insufficient. *American Journal of Epidemiology* 2025;**195**:1472–4. doi:10.1093/aje/kwaf116

6 Van Der Bij S, Koffijberg H, Lenters V, *et al.* Lung Cancer Risk at Low Cumulative Asbestos Exposure: Meta-Regression of the Exposure–Response Relationship. *Cancer Causes & Control* 2013;**24**:1–12. doi:10.1007/s10552-012-0107-7

7 Debray TPA, Moons KGM, Abo-Zaid GMA, *et al.* Individual Participant Data Meta-Analysis for a Binary Outcome: One-Stage or Two-Stage?. *PLoS ONE* 2013;**8**:e60650. doi:10.1371/journal.pone.0060650

8 Stucker I. Genetic polymorphisms of glutathione S-transferases as modulators of lung cancer susceptibility. *Carcinogenesis* 2002;**23**:1475–81. doi:10.1093/carcin/23.9.1475

9 Ilar A, Plato N, Lewné M, *et al.* Occupational exposure to diesel motor exhaust and risk of lung cancer by histological subtype: a population-based case-control study in Swedish men. *European Journal of Epidemiology* 2017;**32**:711–9. doi:10.1007/s10654-017-0268-5

10 Brenner DR, Hung RJ, Tsao M-S, *et al.* Lung cancer risk in never-smokers: a population-based case-control study of epidemiologic risk factors. *BMC Cancer* 2010;**10**:285. doi:10.1186/1471-2407-10-285

11 Beulens JWJ, Monninkhof EM, Verschuren WMM, *et al.* Cohort Profile: The EPIC-NL study. *International Journal of Epidemiology* 2009;**39**:1170–8. doi:10.1093/ije/dyp217

12 Riley RD, Debray TPA. The One-Stage Approach to IPD Meta-Analysis. In: Riley RD, Tierney JF, Stewart LA, eds. *Individual Participant Data Meta-Analysis*. 2021. 127–62. doi:10.1002/9781119333784.ch6

13 Kerr S, Greenland S, Jeffrey K, *et al.* Understanding and reporting odds ratios as rate-ratio estimates in case-control studies. *Journal of Global Health* 2023;**13**:4101. doi:10.7189/jogh.13.04101

14 Greenland S, Thomas DC. On the Need for the Rare Disease Assumption in Case-Control Studies. *American Journal of Epidemiology* 1982;**116**:547–53. doi:10.1093/oxfordjournals.aje.a113439

15 Vandenbroucke JP, Pearce N. Case-Control Studies: Basic Concepts. *International Journal of Epidemiology* 2012;**41**:1480–9. doi:10.1093/ije/dys147

16 Symons M, Moore D. Hazard Rate Ratio and Prospective Epidemiological Studies. *Journal of Clinical Epidemiology* 2002;**55**:893–9. doi:10.1016/S0895-4356(02)00443-2

17 Riley RD, Debray TP, Fisher D, *et al.* Individual Participant Data Meta-analysis to Examine Interactions between Treatment Effect and Participant-level Covariates: Statistical Recommendations for Conduct and Planning. *Statistics in Medicine* 2020;**39**:2115–37. doi:10.1002/sim.8516

18 Lenters V, Vermeulen R, Dogger S, *et al.* A Meta-analysis of Asbestos and Lung Cancer: Is Better Quality Exposure Assessment Associated with Steeper Slopes of the Exposure–Response Relationships?. *Environmental Health Perspectives* 2011;**119**:1547–55. doi:10.1289/ehp.1002879

19 Clin B, Morlais F, Launoy G, *et al.* Cancer incidence within a cohort occupationally exposed to asbestos: a study of dose-response relationships. *Occupational and Environmental Medicine* 2011;**68**:832–6. doi:10.1136/oem.2010.059790

20 Courtice MN, Wang X, Lin S, *et al.* Exposure-response estimate for lung cancer and asbestosis in a predominantly chrysotile-exposed Chinese factory cohort. *American Journal of Industrial Medicine* 2016;**59**:369–78. doi:10.1002/ajim.22579

21 Wang X, Yano E, Lin S, *et al.* Cancer Mortality in Chinese Chrysotile Asbestos Miners: Exposure-Response Relationships. *PLoS ONE* 2013;**8**:e71899. doi:10.1371/journal.pone.0071899

22 Pira E, Romano C, Donato F, *et al.* Mortality from cancer and other causes among Italian chrysotile asbestos miners. *Occupational and Environmental Medicine* 2017;**74**:558–63. doi:10.1136/oemed-2016-103673

23 Larson TC, Antao VC, Bove FJ. Vermiculite Worker Mortality: Estimated Effects of Occupational Exposure to Libby Amphibole. *Journal of Occupational & Environmental Medicine* 2010;**52**:555–60. doi:10.1097/jom.0b013e3181dc6d45

24 Heederik D, Santonen T. Opinion on Scientific Evaluation of Occupational Exposure Limits for Asbestos. 2021.

https://echa.europa.eu/documents/10162/7937606/OEL_asbestos_Final_Opinion_en.pdf/

(accessed 11 Nov 2024).

25 Lidell F, McDonald A, McDonald J. The 1891-1920 birth cohort of Quebec Chrysotile miners and millers: Development from 1904 and mortality to 1992. *The Annals of Occupational Hygiene* 1997;**41**:13–36. doi:10.1016/s0003-4878(96)00044-0

26 McDonald AD, Fry JS, Woolley AJ, *et al.* Dust exposure and mortality in an American chrysotile asbestos friction products plant. *Occupational and Environmental Medicine* 1984;**41**:151–7. doi:10.1136/oem.41.2.151

27 Hein MJ, Stayner LT, Lehman E, *et al.* Follow-up study of chrysotile textile workers: cohort mortality and exposure-response. *Occupational and Environmental Medicine* 2007;**64**:616–25. doi:10.1136/oem.2006.031005

28 Loomis D, Dement JM, Wolf SH, *et al.* Lung cancer mortality and fibre exposures among North Carolina asbestos textile workers. *Occupational and Environmental Medicine* 2009;**66**:535–42. doi:10.1136/oem.2008.044362

29 Berry G, Klerk NH de, Reid A, *et al.* Malignant pleural and peritoneal mesotheliomas in former miners and millers of crocidolite at Wittenoom, Western Australia. *Occupational and Environmental Medicine* 2004;**61**:e14–4. doi:10.1136/oem.2003.008128

30 Seidman H, Selikoff IJ, Gelb SK. Mortality experience of amosite asbestos factory workers: Dose-response relationships 5 to 40 years after onset of short-term work exposure. *American Journal of Industrial Medicine* 1986;**10**:479–514. doi:10.1002/ajim.4700100506

31 Levin JL, McLarty JW, Hurst GA, *et al.* Tyler asbestos workers: mortality experience in a cohort exposed to amosite. *Occupational and Environmental Medicine* 1998;**55**:155–60. doi:10.1136/oem.55.3.155

32 Berry G, Newhouse ML. Mortality of workers manufacturing friction materials using asbestos. *Occupational and Environmental Medicine* 1983;**40**:1–7. doi:10.1136/oem.40.1.1

33 Finkelstein MM. Mortality Among Employees of an Ontario Asbestos-Cement Factory. *American Review of Respiratory Disease* 1984;**129**:754–61. doi:10.1164/arrd.1984.129.5.754

34 Hughes JM, Weill H, Hammad YY. Mortality of workers employed in two asbestos cement manufacturing plants. *Occupational and Environmental Medicine* 1987;**44**:161–74. doi:10.1136/oem.44.3.161

35 Albin M, Jakobsson K, Attewell R, *et al.* Mortality and cancer morbidity in cohorts of asbestos cement workers and referents. *Occupational and Environmental Medicine* 1990;**47**:602–10. doi:10.1136/oem.47.9.602

36 Lacquet LM, van der Linden L, Lepoutre J. Roentgenographic Lung Changes, Asbestosis and Mortality in a Belgian Asbestos-Cement Factory. *IARC scientific publications* 1980;:783–93.

37 Enterline PE, Hartley J, Henderson V. Asbestos and cancer: a cohort followed up to death. *Occupational and Environmental Medicine* 1987;**44**:396–401. doi:10.1136/oem.44.6.396

38 Selikoff IJ, Seidman H. Asbestos-Associated Deaths among Insulation Workers in the United States and Canada, 1967-1987. *Annals of the New York Academy of Sciences* 1991;**643**:1–14. doi:10.1111/j.1749-6632.1991.tb24439.x

39 McDonald AD, Fry JS, Woolley AJ, *et al.* Dust exposure and mortality in an American factory using chrysotile, amosite, and crocidolite in mainly textile manufacture. *Occupational and Environmental Medicine* 1983;**40**:368–74. doi:10.1136/oem.40.4.368

40 Peto J, Doll R, Hermon C, *et al.* Relationship of Mortality to Measures of Environmental Asbestos Pollution in an Asbestos Textile Factory. *The Annals of Occupational Hygiene* 1985;**29**:305–55. doi:10.1093/annhyg/29.3.305

41 Olsson AC, Vermeulen R, Schüz J, *et al.* Exposure–Response Analyses of Asbestos and Lung Cancer Subtypes in a Pooled Analysis of Case–Control Studies:. *Epidemiology* 2017;**28**:288–99. doi:10.1097/EDE.0000000000000604

42 Bruske-Hohlfeld I, Muhner M, Pohlabeln H, *et al.* Occupational Lung Cancer Risk for Men in Germany: Results from a Pooled Case-Control Study. *American Journal of Epidemiology* 2000;**151**:384–95. doi:10.1093/oxfordjournals.aje.a010218

43 Felicitas López-Cima M, González-Arriaga P, García-Castro L, *et al.* Polymorphisms in XPC, XPD, XRCC1, and XRCC3 DNA repair genes and lung cancer risk in a population of Northern Spain. *BMC Cancer* 2007;**7**:162. doi:10.1186/1471-2407-7-162

44 Consonni D, De Matteis S, Lubin JH, *et al.* Lung Cancer and Occupation in a Population-based Case-Control Study. *American Journal of Epidemiology* 2010;**171**:323–33. doi:10.1093/aje/kwp391

45 Jockel K. Occupational risk factors for lung cancer: a case-control study in West Germany. *International Journal of Epidemiology* 1998;**27**:549–60. doi:10.1093/ije/27.4.549

46 Papadopoulos A, Guida F, Cénée S, *et al.* Cigarette smoking and lung cancer in women: Results of the French ICARE case-control study. *Lung Cancer* 2011;**74**:369–77. doi:10.1016/j.lungcan.2011.04.013

47 Brennan P, Crispo A, Zaridze D, *et al.* High Cumulative Risk of Lung Cancer Death among Smokers and Nonsmokers in Central and Eastern Europe. *American Journal of Epidemiology* 2006;**164**:1233–41. doi:10.1093/aje/kwj340

48 Carel R, Olsson AC, Zaridze D, *et al.* Occupational exposure to asbestos and man-made vitreous fibres and risk of lung cancer: a multicentre case-control study in Europe. *Occupational and Environmental Medicine* 2006;**64**:502–8. doi:10.1136/oem.2006.027748

49 Ramanakumar AV, Parent M-E, Siemiatycki J. Risk of Lung Cancer from Residential Heating and Cooking Fuels in Montreal, Canada. *American Journal of Epidemiology* 2007;**165**:634–42. doi:10.1093/aje/kwk117

50 Feyler A, Voho A, Bouchardy C, *et al.* Point: Myeloperoxidase -463G ⟶ A Polymorphism and Lung Cancer Risk. *Cancer Epidemiology, Biomarkers & Prevention* 2002;**11**:1550–4.

51 Fortes C, Forastiere F, Farchi S, *et al.* The Protective Effect of the Mediterranean Diet on Lung Cancer. *Nutrition and Cancer* 2003;**46**:30–7. doi:10.1207/s15327914nc4601_04

52 Richiardi L, Boffetta P, Simonato L, *et al.* Occupational Risk Factors for Lung Cancer in Men and Women: A Population-Based Case-Control Study in Italy. *Cancer Causes & Control* 2004;**15**:285–94. doi:10.1023/b:caco.0000024223.91059.ed

53 Kazma R, Babron M-C, Gaborieau V, *et al.* Lung cancer and DNA repair genes: multilevel association analysis from the International Lung Cancer Consortium. *Carcinogenesis* 2012;**33**:1059–64. doi:10.1093/carcin/bgs116
